# Analysis of electronic health record data of hepatitis B virus (HBV) patients in primary care: hepatocellular carcinoma (HCC) risk associated with socioeconomic deprivation and reduced by statins

**DOI:** 10.1101/2022.09.01.22279481

**Authors:** Cori Campbell, Tingyan Wang, Iain Gillespie, Eleanor Barnes, Philippa C Matthews

## Abstract

**Background:** We set out to characterise chronic Hepatitis B (CHB) in the primary care population in England and investigate risk factors for progression to hepatocellular carcinoma (HCC).

**Methods:** We identified 8039 individuals with CHB in individuals aged ≥18 years between 1999-2019 in the English primary care database QResearch. HCC risk factors were investigated using Cox proportional hazards modelling.

**Findings:** Most of those living with CHB were males (60%) of non-White ethnicity (>70%), and a high proportion were in the most deprived Townsend deprivation quintile (44%). Among 7029 individuals with longitudinal data, 161 HCC cases occurred. Increased HCC hazards significantly associated with male sex (adjusted hazards ratio (aHR) 3.44, 95% Confidence Interval (95CI) 2.07-5.73), older age (for age groups 56-55 and ≥66 years of age, compared to 26-35 years, aHRs 7.52 (95CI 4.14-13.67) and11.89 (95CI 6.26-22.60) respectively), socioeconomic deprivation (aHR for fifth Townsend deprivation quintile 1.69, 95CI 1.01-2.84, compared to third), Caribbean ethnicity (aHR 3.32, 95CI 1.43-7.71, compared to White ethnicity), ascites (aHR 1.85, 95CI 1.02-3.36), cirrhosis (aHR 6.52, 95CI 4.54-9.37) and peptic ulcer disease (aHR 2.20, 95CI 1.39-3.49). Reduced HCC hazards were associated with statin use (aHR 0.47, 95CI 0.22-0.99).

**Interpretation:** Targeting resources at vulnerable groups, and addressing modifiable risk factors is essential to improve CHB outcomes, and to support progress towards international goals for the elimination of hepatitis infection as a public health threat.

**Funding:** Wellcome (grant ref 110110/Z/15/Z), UCLH NIHR Biomedical Research Centre, Nuffield Department of Medicine, University of Oxford, GlaxoSmithKline, NIHR Health Informatics Collaborative, Cancer Research UK.

**Research in context:** *Evidence before this study:* THE CHB population in England has not been well described. Hepatitis B virus (HBV) reports from the UK Health Security Agency (UHKSA) have not previously reported chronic HBV (CHB) prevalence stratified by relevant subgroups, including ethnicity and socioeconomic status. The burdens of comorbid diseases in this population have also not been characterised. Furthermore, risk factors for the progression of CHB to hepatocellular carcinoma (HCC) have previously been identified largely in homogenous patient samples which may not be widely generalisable. Therefore, risk factors identified in previously published studies require validation in diverse multi-ethnic cohorts. Characterisation of CHB and investigation of novel risk factors for HCC is warranted in a large data source which contains parameters for a large percentage of the population which are collected in a systematic and wide-scale manner in order to improve generalisation of findings.

*Added value of this study:* We have characterised the largest cohort of CHB individuals in the UK to date, using the QResearch primary care electronic health record database, and describing the demographics and burdens of comorbid disease in the population. This is novel and has not previously been done in a large socioeconomically and ethnically diverse patient sample. We have also analysed risk factors for HCC in the cohort, both validating previously reported factors and investigating novel factors.

*Implications of all the available evidence:* The findings of this study have important implications for CHB prevention, clinical management, and resource planning. Our detailed description of the demographics and disease profile of the CHB population in the UK may facilitate the targeting of health and prevention resources. Findings concerning HCC risk factors have implications for the clinical management of CHB in order to reduce the risk of progression to HCC.

## INTRODUCTORY STATEMENT

The 2022 Global Burden of Disease analysis of hepatitis B virus (HBV) epidemiology estimates that >300 million people live with chronic infection worldwide (1). Through its progression to inflammatory liver disease, cirrhosis and primary liver cancer (hepatocellular carcinoma (HCC)), chronic hepatitis B (CHB) accounts for substantial morbidity and mortality, being the leading global cause of HCC death (2), and the third largest cause of cirrhosis death (1). Age-standardised death rates have remained constant or increased for HCC and cirrhosis, respectively, and the total number of HBV-attributable deaths has increased worldwide over recent decades (3). International targets calling for the elimination of HBV infection as a public health threat by the year 2030 have been set (4), with recent recognition and investment into the early detection and treatment of HCC to reduce morbidity and mortality (5). Meeting elimination targets relies on a clear understanding of the epidemiology of infection and associated liver disease in order to target resources and interventions to high-risk groups, and to benchmark progress.

CHB prevalence has not been robustly estimated in many settings, including the UK (6), and groups at the highest risk of morbidity and mortality have not been well characterised. Furthermore, even in well-defined CHB populations, treatment coverage and eligibility are often unreported. Regional HBV reports from UK public health services (UK Health Security Agency, previously Public Health England) have included neither overall estimates of the proportion of CHB individuals receiving antiviral treatment, nor estimates stratified by relevant subgroups such as age, sex and ethnicity (6–10).

There has been increasing interest in identifying risk factors for progression of CHB to cirrhosis, HCC and other endpoints (11). Age, sex, viral load and viral genotype are recognised determinants of HCC risk (12–18), and recent studies have reported associations between HCC and various comorbidities, including type 2 diabetes mellitus (T2DM) and hypertension (11). Existing risk scores, including PAGE-B (19), REACH-B (20, 21), GAG-HCC (22, 23) and CU-HCC (24–26), use such risk factors to predict HCC. However, few cohorts have been characterised in European countries and/or in ethnically diverse populations, to validate or inform scoring approaches. Studies based on electronic health records (EHRs) enable characterisation of large retrospective cohorts, thus enhancing statistical power, and identifying a study sample that is more representative of the whole disease population compared to clinical trials. Such databases often have longitudinal follow-up, with exposures and outcomes ascertained over time. EHR databases can often be linked to other registries (such as national cancer registries and vital statistics), allowing for identification of relevant endpoints.

Given the substantial evidence gaps concerning HBV epidemiology, disease burden and risk factors for progression to HCC, we set out with two aims (i) to characterise a CHB cohort collating data from a large-scale primary care database in England (27), and (ii) to investigate risk factors for progression to HCC.

## PATIENTS AND METHODS

### Data source and study population/design

We used data from the England primary care database QResearch (version 45), which contains >35 million patient records from >1800 individual practices (28). QResearch was established in 2002 and contains anonymised individual-level patient EHRs. Data are collected prospectively and are linked to hospital episode statistics (HES), National Cancer Registration Analysis Service (NCRAS) and Office for National Statistics (ONS) mortality data.

We first identified individuals from the QResearch database between 01 January 1999 and 31 December 2019 who were age ≥18 years and had a record of CHB based on a diagnostic Systemised Nomenclature of Medicine (SNOMED)/Read or International Classification of Disease (ICD) code, or who had a history of ≥1 hepatitis B surface antigen (HBsAg) or HBV DNA viral load (VL) measurement. From this sample we then selected patients for inclusion in our cohort who (at any time between 01 January 1999 and 31 December 2019) were aged ≥18 years, and has confirmed CHB. Patients were identified as having CHB if they met any one of the following conditions: (i) record of a diagnostic SNOMED/Read code indicating CHB; (ii) record of a diagnostic ICD-9 or −10 code indicating CHB, and/or; (iii) Presence of HBsAg or VL on at least two recordings ≥6 months apart (**Supplementary Figure 1**).

### Covariate selection and ascertainment

We identified relevant covariates for extraction *a priori* based on previous literature (11,19,21–26,29) and clinical relevance. Lifestyle factors, demographics and relevant numeric biomarkers were characterised from relevant SNOMED/Read codes. Comorbidities were characterised from relevant SNOMED/Read and ICD-9 and −10 codes. Ethnicity is categorised in QResearch as per 2011 census categories (30). Townsend Deprivation quintile is available as a measure of socioeconomic status in QResearch. Body mass index (BMI, kg/m^2^) was categorised (underweight, <18.5 kg/m^2^; healthy weight, 18.5-2.49 kg/m^2^; overweight, 25.0-29.9 kg/m^2^; obese, ≥30 kg/m^2^) based on World Health Organization (WHO) categories (31). Covariate measurements made within ±3 years of earliest CHB diagnosis and before HCC diagnosis were used as proxy baseline measurements. Where patients had >1 measurement taken within 3 years of the earliest CHB diagnosis, measurements taken closest to diagnosis date were used.

### Outcome ascertainment

Our primary endpoint of interest was HCC. HCC was ascertained via identification of patients with relevant SNOMED/Read or ICD codes corresponding to HCC, and by linkage of the cohort to National Cancer Registry data (32, 33). In order to maximise outcome ascertainment, we used a broad definition for HCC including multiple codes relevant to HCC (**Supplementary Table 1**). We performed sensitivity analysis (further details below) whereby all patients with non-HCC neoplasms were excluded, to investigate robustness of main analysis using our broad HCC definition. A tabulation of HCC cases across source of diagnosis is depicted in **Supplementary Table 2**.

### Follow-up

Earliest date of CHB diagnosis was regarded as cohort entry and initiation of follow-up for each patient. For patients who developed HCC, date of HCC diagnosis was regarded as the end of follow-up. For patients who did not develop HCC (i.e., patients who were censored), follow-up ended at patient cohort exit date (either due to leaving their general practice and switching to a practice which does not contribute to QResearch, or death) or 31 December 2019, whichever occurred earlier. Patients in whom database exit date preceded or was equal to first recorded CHB diagnosis date (n = 1010) whereby follow-up time 0≤ years were excluded from longitudinal analysis.

In some patients, HCC diagnosis date or cohort exit date preceded or was equal to CHB diagnosis date (**Supplementary Table 3**). Data from these patients were excluded from analyses of HCC risk factors.

### Statistical analysis

Statistical analyses were carried out in R (version 4.1.0). Baseline characteristics were summarised for all CHB patients (regardless of length of follow-up) using descriptive statistics. Means and standard deviations (SDs) or medians and interquartile ranges (IQRs) were presented for continuous measures, and were compared using *t* or Wilcoxon rank-sum tests, respectively. Patient counts and percentages were presented for continuous and binary variables, and were compared using chi-squared or Fisher’s exact tests.

Univariable and multivariable Cox proportional hazards models were used to investigate risk factors for progression of CHB to HCC. Variables were included in the multivariable model based on significance of univariable associations (where *P*≤ 0.1) and/or based on biological/clinical relevance and previous literature (11,19,21–26,29). A previous meta-analysis we undertook to investigate risk factors for HCC in CHB was also used to inform variable selection (11).

Continuous laboratory parameters which were right-skewed were transformed with a natural logarithm for inclusion in multivariable models. Laboratory parameters were divided into quintiles for inclusion in multivariable models. Means and SDs for log AST, log alanine transaminase (ALT) and platelet count (Plt) quintiles are presented in supplement (**Supplementary Table 4**). Hazard ratios and 95% confidence intervals (95% CI) were reported for Cox proportional hazards model outputs. Analysis on the imputed dataset was used for main models.

### Handling of missing data

Values were missing for Townsend Deprivation Quintile, ethnicity, alcohol consumption, cigarette consumption, BMI, Plt, ALT measurement, aspartate transaminase (AST) measurement, Hepatitis B surface antigen (HBsAg), and HBV viral load (VL). Missing data are described further in **Supplementary Table 5**.

Multiple imputation by chained equations (MICE) was used to impute missing data across patient characteristics. The assumption of missing at random was made for imputed variables. This is in accordance with previous handling of missing data in cohorts utilising QResearch data (34–37), and current recommendations for imputation of missing data (38). Characteristics with >90% missingness were not imputed. Ten imputed datasets were generated, and results from univariable and multivariable Cox proportional hazards models from each dataset were pooled according to Rubin’s rules (39, 40).

### Sensitivity analyses

To test robustness of our main model, three sensitivity analyses were performed (**Supplementary Table 6**), as follows (i) main results model fit to complete-case cohort subset (i.e. the subset of patients with completeness for all variables); (ii) exclusion of patients with history of non-HCC neoplasms (presented in **Supplementary Table 7**) to control for unmeasured outcome misclassification whereby secondary liver cancer has been misclassified as primary HCC, and; (iii) addition of ALT, AST and Plt in the main model fit to the imputed dataset, as the percentage of missingness in these exposures was too high for them to be included in main analysis.

## RESULTS

### Individuals with CHB are under-represented in primary care records

We identified 8039 CHB patients in the QResearch database between 1999 and 2019 from a database-wide denominator of ∼35 million individuals, which translates to a CHB prevalence of 0.023%. This is an order of magnitude less than current estimates for CHB prevalence in the UK, of 0.27% to 0.73% (41) Thus we have only had access to a dataset representing a fraction of all CHB cases. This discrepancy highlights under-diagnosis of HBV infection, and poor linkage of data between specialist care services and primary care.

### The majority of patients with CHB were male, of non-white ethnicity, in the fourth and fifth (most deprived) Townsend deprivation quintiles, and not receiving treatment

Most of the adults represented in this study were identified by a SNOMED/Read or ICD CHB diagnostic code (7856/8039, 97.7%), with a remaining 2.3% (252/8039) identified by HBsAg and/or VL measurements (in the absence of a diagnostic code) (**Table 1**). Median follow-up duration was 3.87 years (IQR 6.30 years), with differential follow-up between individuals who developed HCC (median follow-up 1.47 years, IQR 5.13 years) and those who did not (median follow-up duration 3.93 years, IQR 6.28 years). Mean age at baseline was 38.3 years (SD 11.6 years), and at baseline >75% were ≤45 years of age. The majority were male (4856/8039, 60.4%). Black Africans represented 25.4% of individuals, with 12.9% of Chinese ethnicity, 5.9% of Pakistani ethnicity, ≤ 3.0% of Indian ethnicity and 28.4% of White ethnicity (**Table 1**). Proportions of Black and ethnic minorities in our CHB cohort were greater than those in both the wider QResearch database (42) and general English population (43) (**Figure 2**). At baseline, most (88.0%) of patients had no record of antiviral treatment. Within 1, 2, 3 and ≥ 4 years of CHB diagnoses, cumulatively 2,7%, 4.0 %, 5.0% and 10.1% of patients had record of antiviral treatment initiation, respectively.

**Figure 1.**
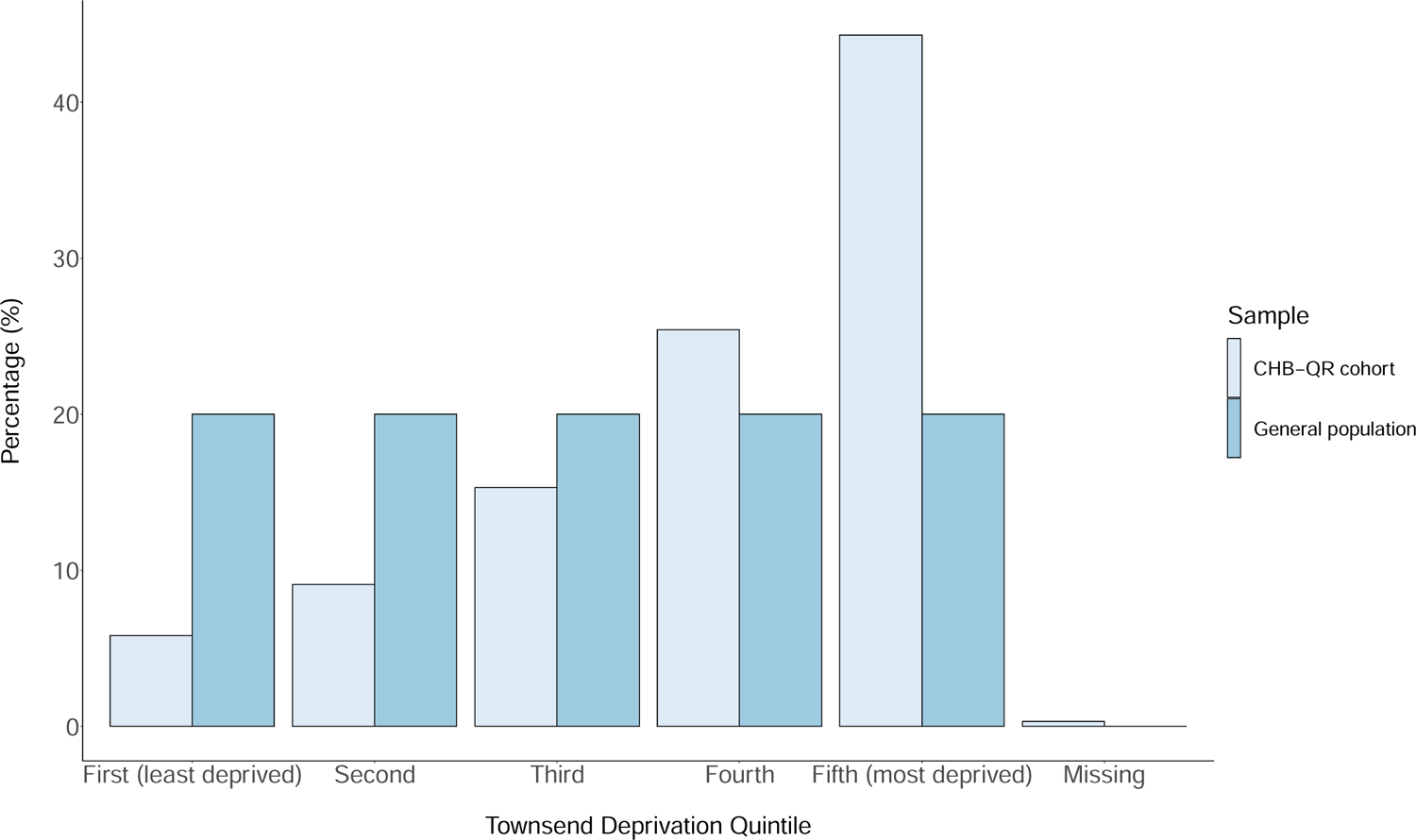
Townsend deprivation quintile breakdown in 8039 adults with chronic hepatitis B virus infection derived from the QResearch primary care database (England) versus the United Kingdom general population.

**Figure 2.**
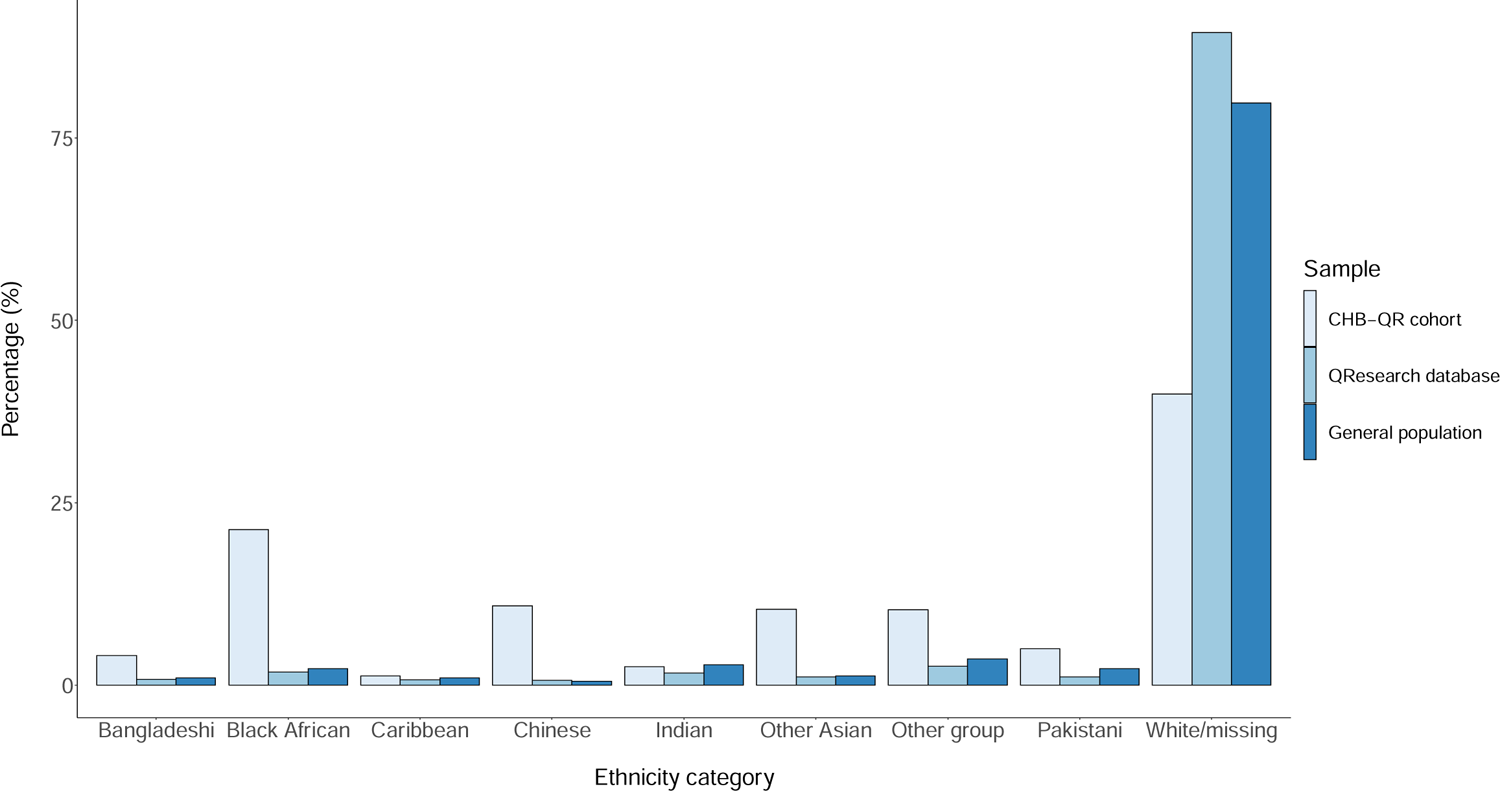
Ethnicity breakdown in 8039 adults in a chronic hepatitis B virus cohort characterised from the QResearch primary care database (England) versus all individuals in the QResearch database (∼35 million), vs. the United Kingdom general population. General population estimates obtained from 2019 estimates from the Office for National Statistics.

**Table 1:**
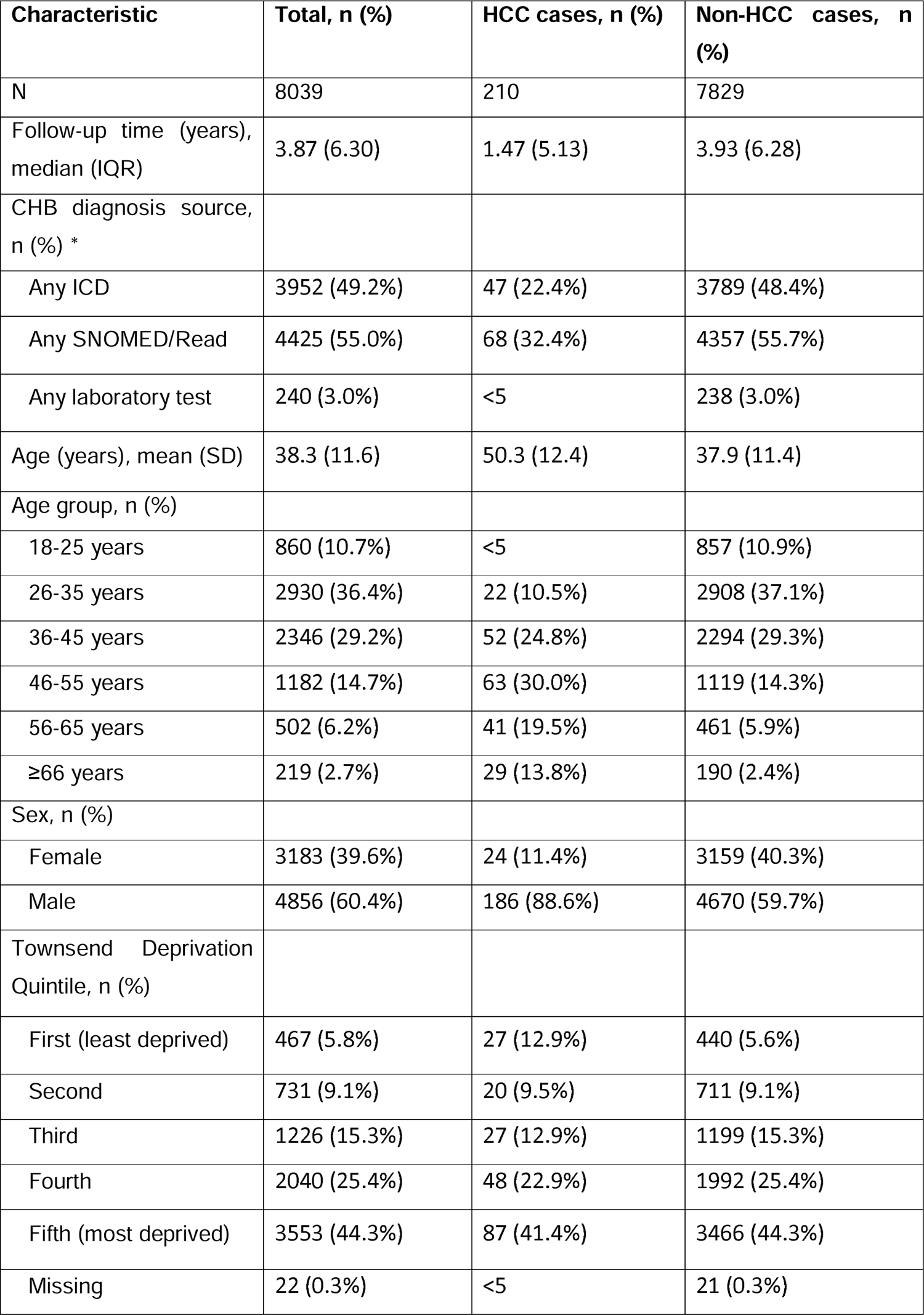
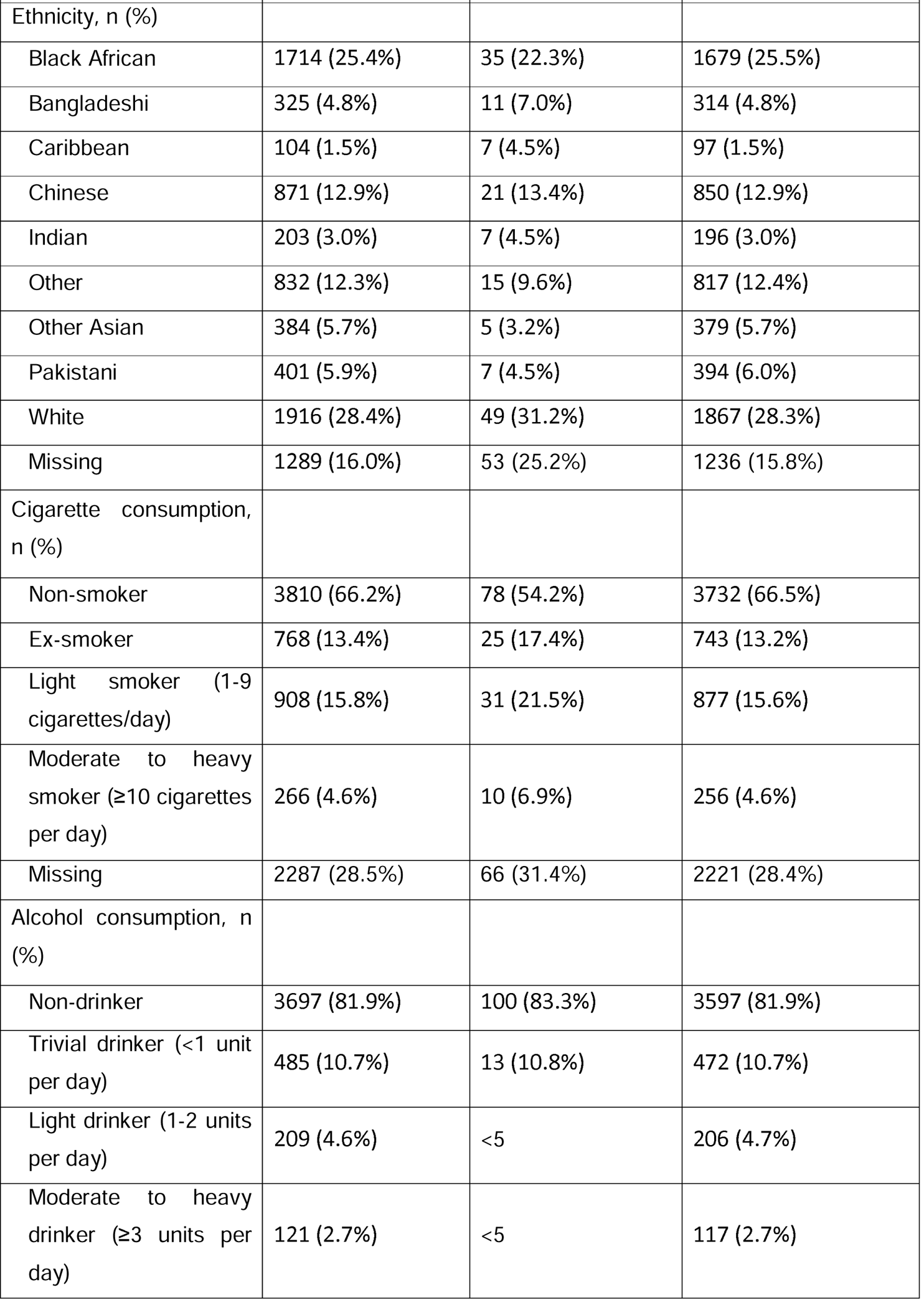
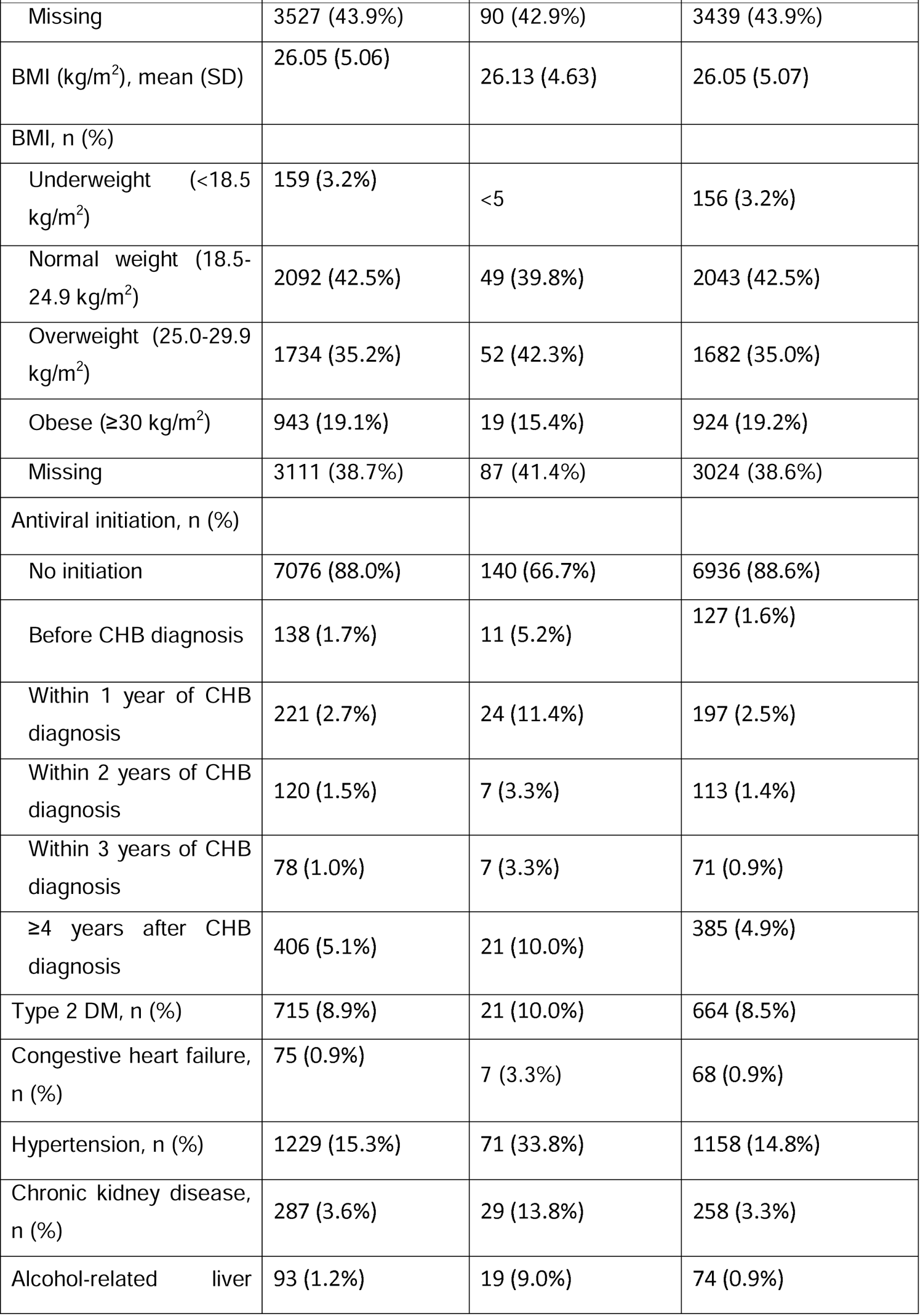
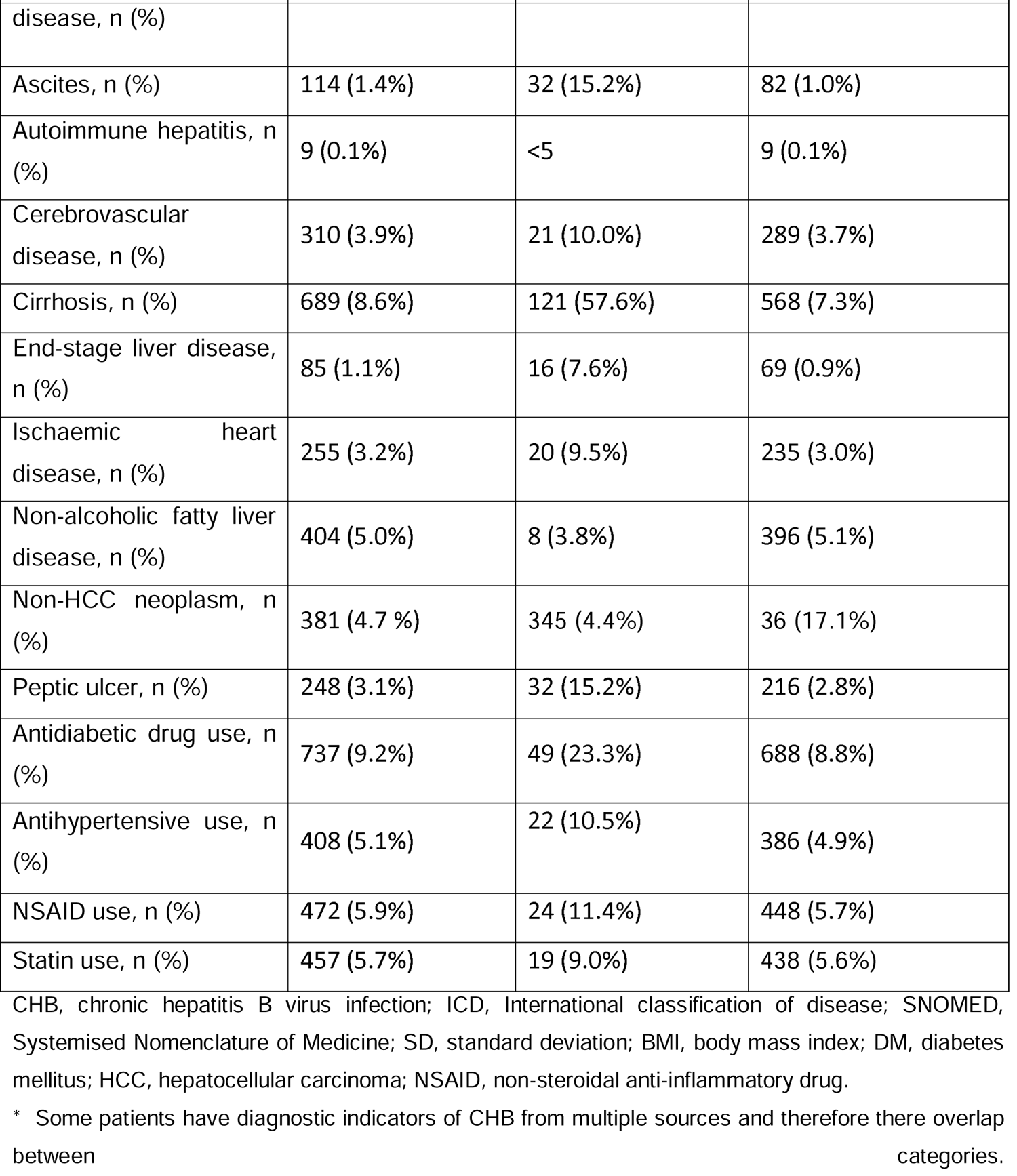
Cohort baseline characteristics

Cohort characteristics stratified by Townsend deprivation quintile are presented in **Table 2**. Age and sex associated significantly with Townsend deprivation quintile whereby mean ages were younger (*P* < 0.001) and proportions of males were higher (*P* = 0.021) in more deprived quintiles. Ethnicity significantly differed across quintiles (*P* < 0.001) whereby the proportions of Bangladeshi and Black African ethnicity patients increased with increasing deprivation and the proportion of White and Chinese patients decreased. Alcohol and cigarette consumption were significantly associated with Townsend deprivation quintile (*P* < 0.001 and *P* = 0.04, respectively), but no obvious trends were apparent. The only comorbidities differing significantly in prevalence across quintiles were cerebrovascular disease, peptic ulcer disease and non-HCC neoplasms. Associations of antidiabetic drug, antihypertensive, NSAID and statin use with Townsend Deprivation Quintile were nonsignificant.

**Table 2.**
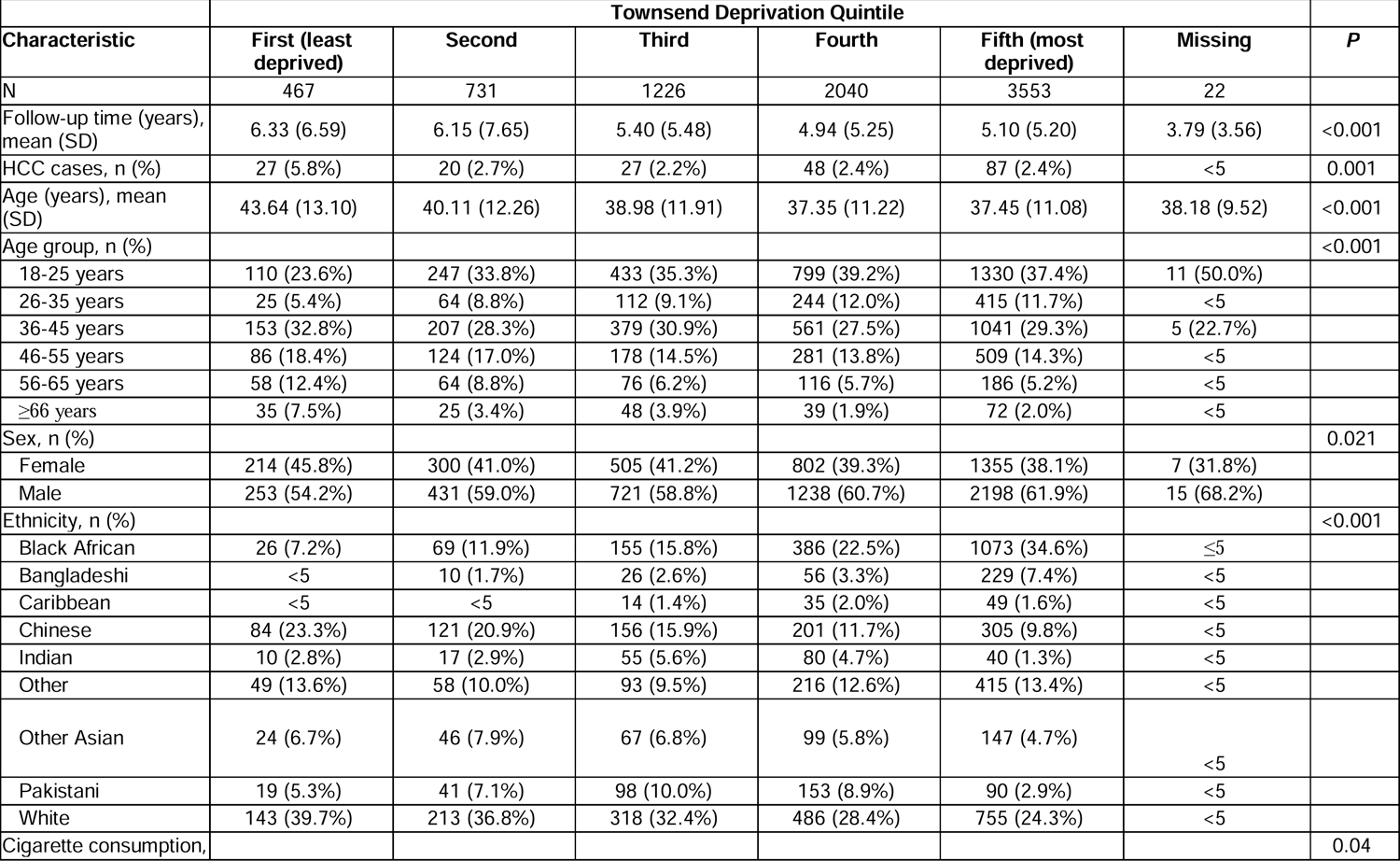
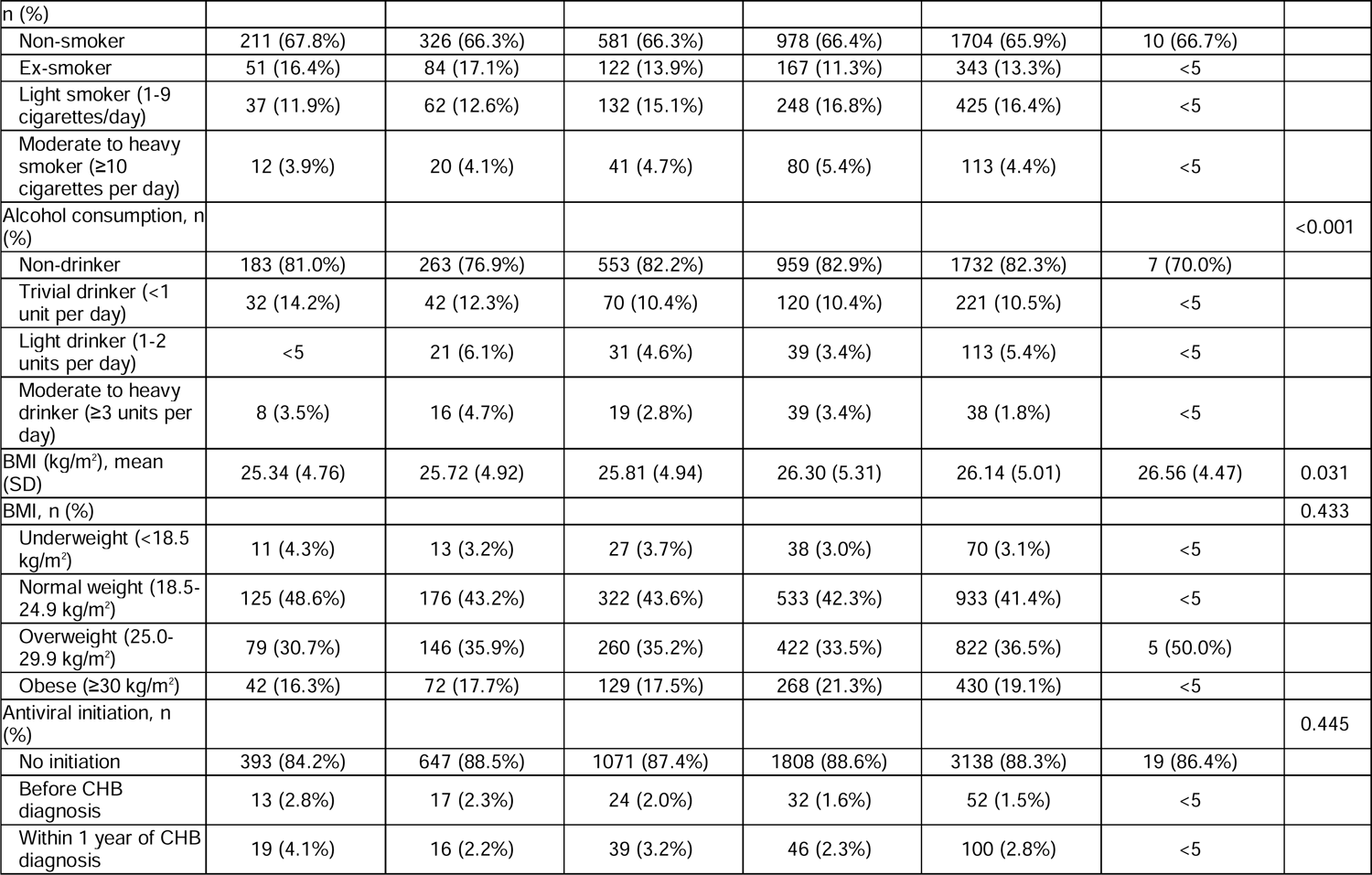
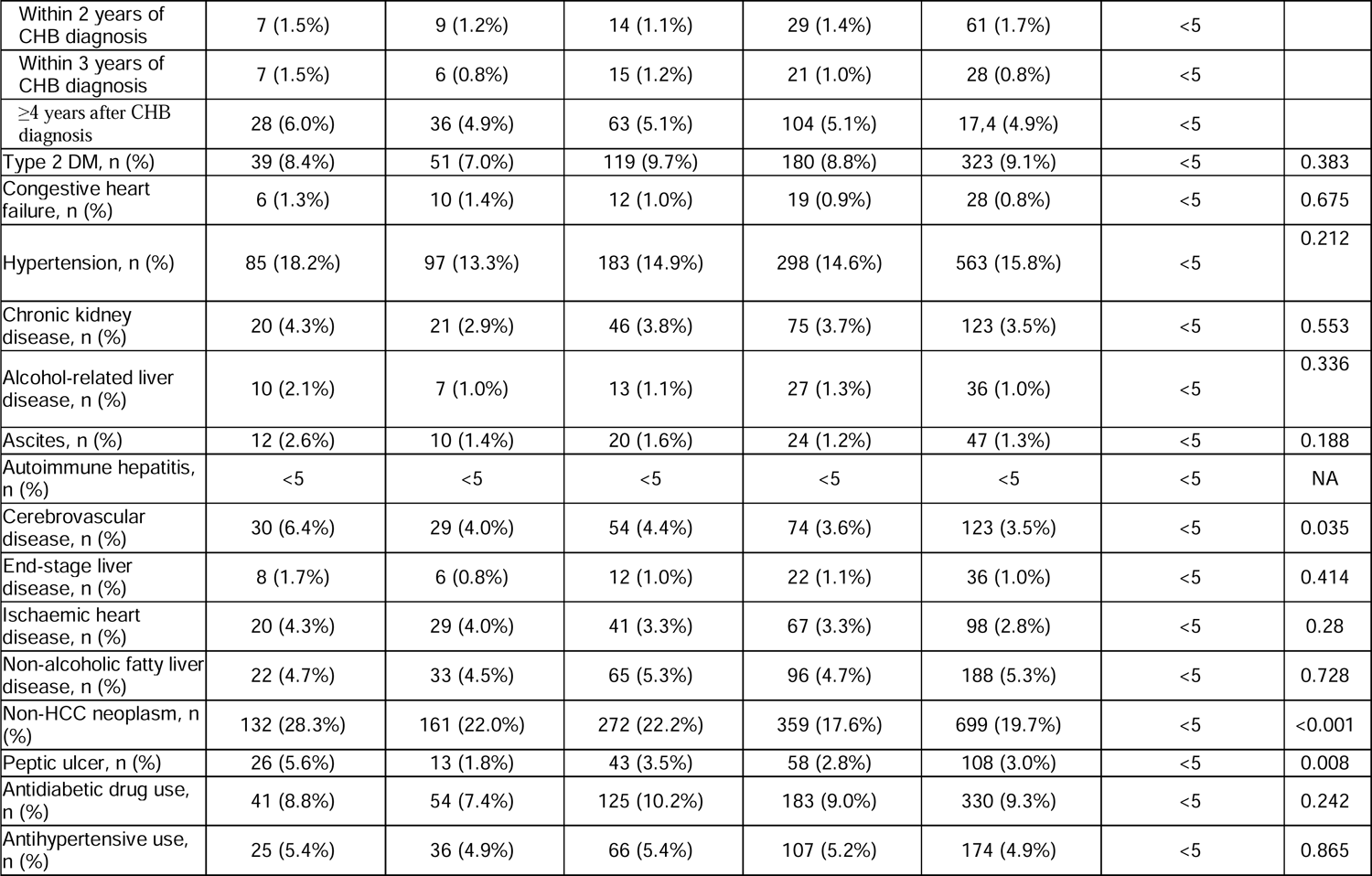
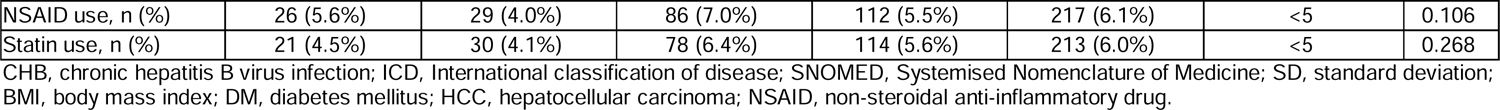
Characteristics of adults with chronic hepatitis B virus infection identified from QResearch primary care database, stratified by socioeconomic status.

### Prevalence of diabetes and hypertension in adults with CHB was higher than in the general population

Baseline prevalence of T2DM and hypertension were 8.9% and 15.3%, respectively. This differs from the prevalence of T2DM and hypertension in the wider QResearch database of <8% and <3%, respectively (44). Prevalence of other comorbidities (including congestive heart failure, chronic kidney disease, alcohol-related liver disease, ascites, autoimmune hepatitis, cerebrovascular disease, end-stage liver disease, ischaemic heart disease, no-alcoholic fatty liver disease and peptic ulcer disease; **Table 1**) ranged from 0.1% to 5%. Baseline cirrhosis is difficult to define from primary care EHR due to lack of relevant laboratory and imaging data, but 8.6% of the cohort had a diagnostic code indicating cirrhosis. Medication use was as follows: antidiabetic drugs (9.2%), antihypertensives (5.1%), non-steroidal anti-inflammatory drugs (NSAIDs) (5.9%) and statins (5.7%). Prevalence of non-HCC neoplasm was 4.7% in the overall cohort. Data for smoking consumption, alcohol consumption and BMI were available for 71.5%, 56.1% and 61.3% of the cohort (**Table 1**).

### Risk factors for HCC included male sex, older age, increased deprivation, Caribbean ethnicity and peptic ulcer disease

Baseline characteristics of the imputed dataset used in analysis of HCC risk factors are presented in **Table 3**. Multivariable Cox proportional hazards models were constructed for 7029 patients in whom 161 HCC cases developed throughout 41 147 person-years of follow-up (**Figure 3**, **Supplementary Table 8**). This translated to an HCC incidence rate of 5.10 cases per 1000 person-years (95% Confidence Interval (95CIs) 4.46 to 5.84).

**Figure 3.**
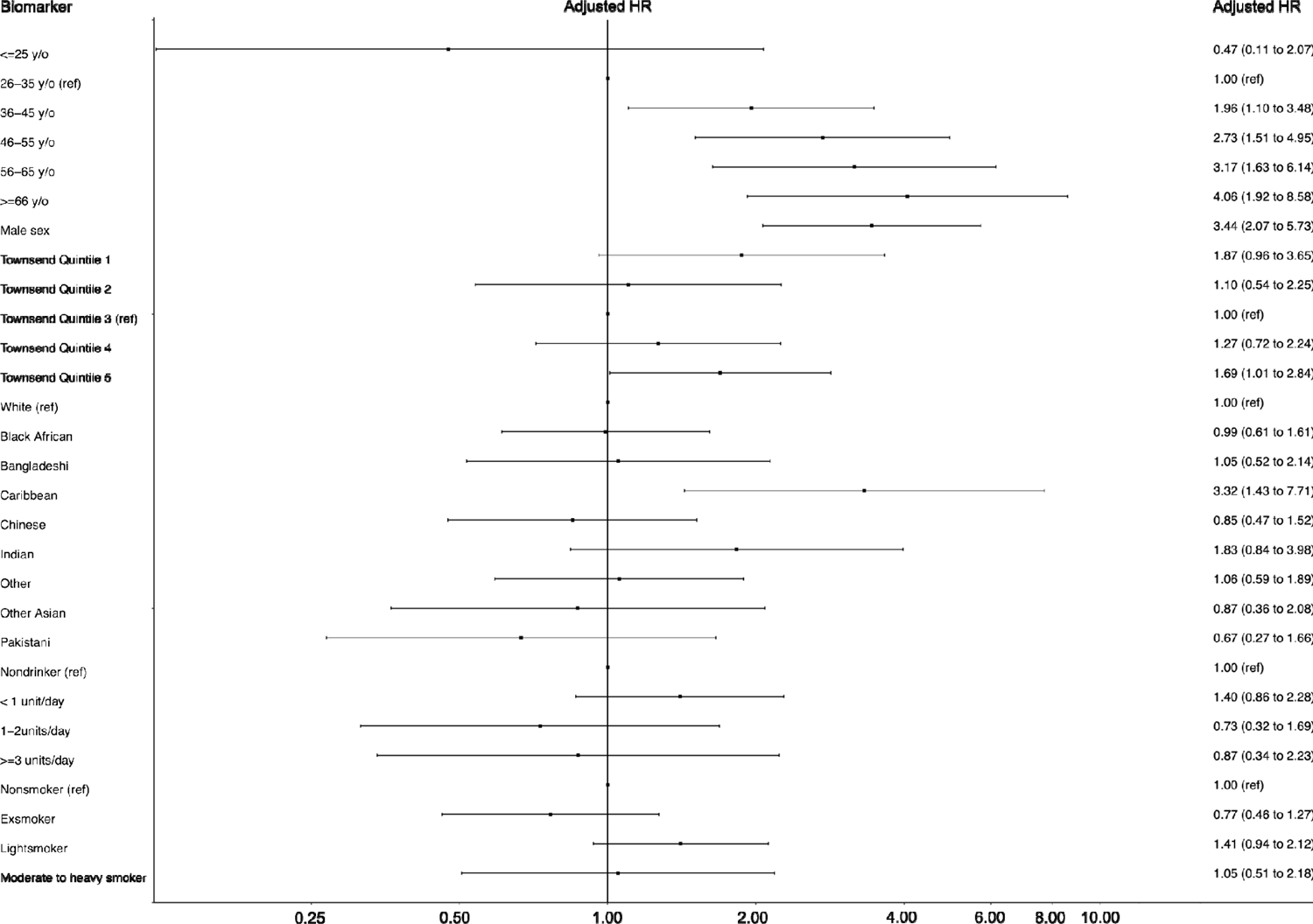
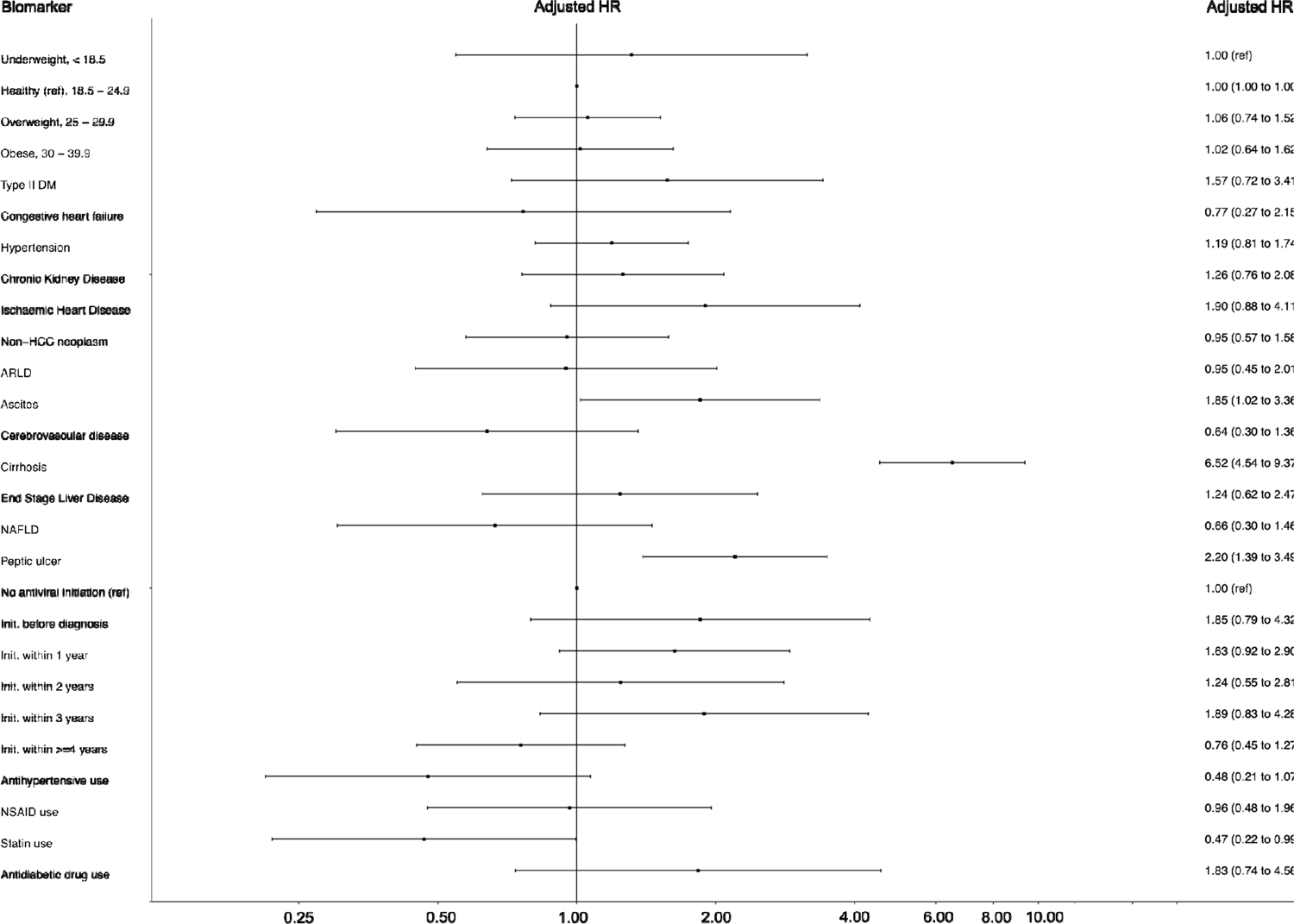
Forest plot for Cox proportional hazards model to identify risks of hepatocellular carcinoma (HCC) in an adult population with chronic Hepatitis B virus infection derived from the QResearch primary care database. Analysed using a dataset generated by multiple imputation with chained equations (n = 7029, HCC cases = 161). First (least deprived) to fifth (most deprived) Townsend Deprivation Quintiles are denoted by SES1-5, respectively.

**Table 3.** Baseline characteristics of complete-case and imputed (by multiple imputation with chained equations) datasets used in Cox proportional hazards models.

| Characteristic | Complete-case dataset, n (%) | Imputed dataset, n (%) |
| --- | --- | --- |
| N | 3648 | 7029* |
| HCC cases, n (%) | 68 (1.9%) | 161 (2.3%) |
| Age group, n (%) |  |  |
| 18-25 years | 292 (8.0) | 659 (9.4) |
| 26-35 years | 1253 (34.3) | 2500 (35.6) |
| 36-45 years | 1126 (30.9) | 2108 (30.0) |
| 46-55 years | 593 (16.3) | 1086 (15.5) |
| 56-65 years | 270 (7.4) | 468 (6.7) |
| ≥66 years | 114 (3.1) | 208 (3.0) |
| Sex, n (%) |  |  |
| Male | 2082 (57.1) | 4186 (59.6) |
| Townsend Deprivation Quintile, n (%) |  |  |
| First (least deprived) | 174 (4.8) | 419 (6.0) |
| Second | 261 (7.2) | 645 (9.2) |
| Third | 523 (14.3) | 1094 (15.6) |
| Fourth | 924 (25.3) | 1781 (25.3) |
| Fifth (most deprived) | 1766 (48.4) | 3090 (44.0) |
| Ethnicity, n (%) |  |  |
| Black African | 956 (26.2) | 1772 (25.2) |
| Bangladeshi | 213 (5.8) | 322 (4.6) |
| Caribbean | 58 (1.6) | 112 (1.6) |
| Chinese | 428 (11.7) | 894 (12.7) |
| Indian | 131 (3.6) | 224 (3.2) |
| Other | 441 (12.1) | 863 (12.3) |
| Other Asian | 194 (5.3) | 397 (5.6) |
| Pakistani | 241 (6.6) | 420 (6.0) |
| White | 986 (27.0) | 2025 (28.8) |
| Cigarette consumption, n (%) |  |  |
| Non-smoker | 2484 (68.1) | 4661 (66.3) |
| Ex-smoker | 480 (13.2) | 924 (13.1) |
| Light smoker (1-9 cigarettes/day) | 531 (14.6) | 1110 (15.8) |
| Moderate to heavy smoker (≥10 cigarettes per day) | 153 (4.2) | 334 (4.8) |
| Alcohol consumption, n (%) |  |  |
| Non-drinker | 3013 (82.6) | 5784 (82.3) |
| Trivial drinker (<1 unit per day) | 371 (10.2) | 737 (10.5) |
| Light drinker (1-2 units per day) | 170 (4.7) | 318 (4.5) |
| Moderate to heavy drinker (≥3 units per day) | 94 (2.6) | 190 (2.7) |
| BMI (kg/m<sup>2</sup>), mean (SD) | 26.14 (5.07) | 26.05 (5.02) |
| BMI, n (%) |  |  |
| Underweight (<18.5 kg/m<sup>2</sup>) | 113 (3.1) | 222 (3.2) |
| Normal weight (18.5-24.9 kg/m<sup>2</sup>) | 1530 (41.9) | 2948 (41.9) |
| Overweight (25.0-29.9 kg/m<sup>2</sup>) | 1292 (35.4) | 2527 (36.0) |
| Obese (≥30 kg/m<sup>2</sup>) | 713 (19.5) | 1332 (19.0) |
| Antiviral initiation, n (%) |  |  |
| No initiation | 3168 (86.8) | 6092 (86.7) |
| Before CHB diagnosis | 67 (1.8) | 121 (1.7) |

| **Characteristic** | **Complete-case dataset, n (%)** | **Imputed dataset, n (%)** |
| --- | --- | --- |
| Within 1 year of CHB diagnosis | 116 (3.2) | 214 (3.0) |
| Within 2 years of CHB diagnosis | 73 (2.0) | 120 (1.7) |
| Within 3 years of CHB diagnosis | 49 (1.3) | 78 (1.1) |
| ≥4 years after CHB diagnosis | 175 (4.8) | 404 (5.7) |
| Type 2 DM, n (%) | 503 (13.8) | 678 (9.6) |
| Congestive heart failure, n (%) | 41 (1.1) | 71 (1.0) |
| Hypertension, n (%) | 754 (20.7) | 1165 (16.6) |
| Chronic kidney disease, n (%) | 175 (4.8) | 271 (3.9) |
| Alcohol-related liver disease, n (%) | 39 (1.1) | 74 (1.1) |
| Ascites, n (%) | 34 (0.9) | 88 (1.3) |
| Autoimmune hepatitis, n (%) | 3 (0.1) | 9 (0.1) |
| Cerebrovascular disease, n (%) | 181 (5.0) | 290 (4.1) |
| Cirrhosis, n (%) | 301 (8.3) | 608 (8.6) |
| End-stage liver disease, n (%) | 28 (0.8) | 78 (1.1) |
| Ischaemic heart disease, n (%) | 143 (3.9) | 229 (3.3) |
| Non-alcoholic fatty liver disease, n (%) | 248 (6.8) | 393 (5.6) |
| Non-HCC neoplasm, n (%) | 167 (4.6) | 340 (4.8) |
| Peptic ulcer, n (%) | 113 (3.1) | 221 (3.1) |
| Antidiabetic drug use, n (%) | 495 (13.6) | 689 (9.8) |
| Antihypertensive use, n (%) | 293 (8.0) | 389 (5.5) |
| NSAID use, n (%) | 331 (9.1) | 446 (6.3) |
| Statin use, n (%) | 342 (9.4) | 443 (6.3) |

CHB, chronic hepatitis B virus infection; ICD, International classification of disease; SNOMED, Systemised Nomenclature of Medicine; IQR, interquartile range; SD, standard deviation; BMI, body mass index; DM, diabetes mellitus; HCC, hepatocellular carcinoma; NSAID, non-steroidal anti-inflammatory drug.

Hazards of HCC were increased in males (adjusted hazards ratio (aHR) 3.44, 95% CI 2.07 to 5.73), with increasing age (aHR for 36-45 years 1.96, 95% CI 1.10 to 3.48; aHR for 46-55 years 2.73, 95% CI 1.51 to 4.95; aHR for 56-65 years 3.17, 95% CI 1.63 to 6.14; aHR for ≥66 years 4.06, 95% CI 1.92 to 8.58, as compared to 26-35 years reference group) and in the fifth deprivation quintile as compared to the third quintile (aHR 1.69, 95% CI 1.01 to 2.84). Hazards of HCC in the Caribbean ethnicity group were higher than those in the White reference group (aHR 3.32, 95% CI 1.43 to 7.71) but did not differ in any other ethnic category. There were no associations between alcohol consumption, cigarette smoking, or BMI with hazards of HCC. As expected, increased hazards of HCC were associated with evidence of advanced liver disease, cirrhosis (aHR 6.52, 95% CI 4.54 to 9.37) and ascites (aHR 1.85, 95% CI 1.02 to 3.36). Interestingly, peptic ulcer disease was also associated with increased HCC hazards (aHR 2.20, 95% CI 1.39 to 3.49). Statin use was associated with reduced hazards of HCC (aHR 0.47, 95% CI 0.22 to 0.99). No other medicines, including antiviral treatment, associated with hazards of HCC. Hazards ratios did not change materially in strength or direction upon sensitivity analysis excluding non-HCC neoplasms or including AST, ALT and Plt at baseline (**Supplementary Table 8**).

### Main model results are robust to complete-case sensitivity analysis

We undertook sensitivity analysis restricted to the subgroup of patients for whom complete data were available (n=3648 patients in whom 68 cases of HCC occurred (**Supplementary Table 9**)). Confidence intervals for all aHRs were more precise in the model from the imputed dataset compared to the complete case analysis due to larger sample size in the former. Hazard ratios did not change materially in strength or direction in sensitivity analysis undertaken to exclude patients with history of non-HCC neoplasms (**Supplementary Table 9**).

## DISCUSSION

### Summary of key findings

This is the largest cohort of CHB individuals characterised in England to date, from either EHR or traditional prospective cohorts. However, our prevalence estimate of 0.023% contrasts with current U.K. prevalence estimates (0.27% to 0.73%) (41), suggesting many cases of HBV infection are not diagnosed and/or not recorded in primary care records. Substantial efforts are needed in clinical, governance and data domains to enhance diagnosis, linkage to care and information linkage, in order to support better individual patient care and to inform public health programmes.

The CHB population we identified was ethnically diverse, with higher proportions of black and ethnic minority individuals than either the wider QResearch database (42) or the general English population (43). Importantly, the CHB population is disproportionately socioeconomically deprived, with substantial burdens of comorbid disease. This highlights a vulnerable population, for whom specific resources should be targeted to redress health inequities. The QResearch database has geographic coverage across England, and therefore our findings should be generalizable to the wider country, and potentially to other settings. However, we recognise the impacts of selection bias and missingness.

We report increased hazards of HCC associated with increasing age, male sex, socioeconomic deprivation, Caribbean ethnicity, severe liver disease (ascites and cirrhosis), and comorbid disease (peptic ulcer disease). Age, sex and T2DM have previously been found to positively associate with HCC risk in CHB (11), however this is the first cohort study to report these associations in an ethnically diverse cohort. It is likely that we are underpowered to detect associations with alcohol and cigarette consumption with HCC risk in our cohort, although smoking was identified as a risk factor on sensitivity analysis.

It is possible that the positive association of peptic ulcer disease with hazards of HCC is confounded by proton pump inhibitor (PPI) administration for peptic ulcer treatment. Information concerning PPI prescription and usage was not available in our QResearch extract, but previous observational studies have reported increased risks of HCC associated with PPIs (45, 46). Pooled risk estimates from meta-analyses have been variable, with some confirming this association (47) and others failing to report associations (48, 49), particularly after restricting to studies of participants with viral hepatitis (49).

### Influence of HBV genotype

HBV genotype is not routinely determined in clinical practice, so is not available from EHR datasets. Genotype is associated with ethnicity (50, 51), and viral genotypes have been associated with both an increased HCC risk (52) and antiviral treatment resistance (53). Therefore, associations of HCC risk with ethnicity and/or socioeconomic status may be driven by HBV genotype. However, the association may also be mediated by unmeasured population genetic factors and/or other exposures (including lifestyle factors). Increased HCC hazards observed in the most deprived Townsend quintile may similarly be confounded by genotype, however, it is more likely the observed association is mediated by socioeconomic factors whereby access to routine care/surveillance is poorer and additional lifestyle or environmental factors which increase HCC risk are more prevalent.

### Drug treatment and HCC development

We report a protective association of statin use with HCC hazards. This association has been reported in previous CHB cohorts (11,54,55), in individuals with predisposing HCC risk factors including cirrhosis and T2DM (56, 57) and in a general patient population (55–57). Further analysis, including potential mediation analysis where data availability allows, is warranted to investigate the mechanisms of this association.

We did not find an association of antiviral treatment with reduced HCC risk, although there is evidence from previous observational and randomised interventional studies that CHB treatment with antiviral nucleoside analogues (NAs) reduces HCC risk (58–60). In our sample, 10.3% of individuals initiated antiviral treatment during follow-up. However, a substantial proportion of treatment data may be missing from primary care records, as this information is typically captured in secondary/tertiary care records where HBV prescribing is based (61). For example, 16% of CHB patients in secondary care were receiving antiviral treatment, based on the NIHR Health Informatics Collaborative (HIC) database (62). Our primary care cohort may systematically differ to samples in clinical trials and hospital-based cohorts whereby only a small proportion of individuals are treatment-eligible in our cohort. Additionally, receipt of antiviral treatment may target those with more severe progression/prognosis compared to untreated individuals at baseline, thus negating any signal for a positive effect. Clinical guidelines recommend criteria for treatment, but it is not possible for us to determine the extent to which the 89.7% of patients with no treatment recorded are treatment ineligible, or are receiving treatment in a secondary/tertiary care setting that is not captured in QResearch data. Therefore, the association of antiviral treatment and HCC risk cannot be confidently determined from this dataset.

### Application of data to HCC risk stratification

HCC risk scores (including PAGE-B (19), REACH-B (20, 21), GAG-HCC (22, 23) and CU-HCC (24–26) utilise various characteristics, including age, sex, and laboratory parameters to predict HCC risk. The utility of existing risk scores in homogenous patient subgroups has been demonstrated (63). Our results highlight the need for validation (and potentially modification) of such scores in heterogenous samples which are ethnically and clinically diverse. Identifying groups who carry a significantly higher risk of developing HCC may inform enhanced screening and support early diagnosis. Determination of modifiable risk factors which influence HCC development should inform interventions, for example a risk associated with smoking should inform specific smoking cessation advice and support.

### Caveats of primary care EHR analysis – missing data

We highlight substantial data missingness due to poor primary care coverage and HBV representation in such data sources. Missingness likely associated with characteristics not measured in our dataset including thereby excluding marginalised subgroups (including undocumented migrants, highly mobile population subgroups, and people who do not speak English), which may further increase the proportion who fall into the most socioeconomically deprived quintiles.

Most patients were identified by coding, with a minority (3.0%) having confirmatory laboratory tests accessible in QResearch. This is unsurprising given that specialist referral is recommended following a positive test result (64) thus second confirmatory HBV lab data (VL and/or HBsAg) would typically be performed in a secondary or tertiary care centre, and these results are not reliably transferred into primary care EHR systems. Efforts to enhance linkage of data between primary and secondary care would be of significant benefit to patient care, and potentially increase the chance of timely diagnosis of liver complications, including HCC, in patients presenting to their primary care practitioner with early symptoms.

Length of follow-up is shorter than anticipated for a chronic disease despite a 20-year study period from 1999 to 2019. This is not uncommon in EHR databases in which there may be time lags between notification of a patient characteristic/disease and input into electronic record systems. Additionally, given the neglected nature of CHB, it is likely that individuals present to primary care late in the infection course and/or diagnoses are primary managed in secondary/tertiary services without linkage to primary care records. Follow-up was differential between individuals who did and did not develop HCC, with shorter median follow-up duration in HCC cases. This may be due to HCC cases only presenting to a GP practice when their liver disease has progressed to a symptomatic, more advanced stage.

### Analytical corrections for missing data

We have undertaken imputation of missing baseline data, in line with previous investigations in QResearch (34–37), but were unable to impute important HBV biomarkers (including VL and HBsAg) as >90% of participants were missing any measurement at baseline. We therefore excluded these variables from analysis. High missingness was observed for additional biomarkers, specifically AST, ALT and Plt, which serve as indicators of liver function and can be used to derive proxy scores for staging fibrosis and cirrhosis. Replication of our present analysis in secondary care datasets is a future opportunity to investigate the utility of these biomarkers as markers of disease endpoints. Similarly, we were unable to time-update our models for changes in alcohol and cigarette consumption throughout follow-up due to lack of repeated measurements (>90% of the cohort had a one-off record for alcohol and smoking). Due to missing data, we were also unable to validate and/or modify such scores. Future investigation in data sources where variables are more complete and remeasurement is more comprehensive is warranted to enable these analytical improvements, and allow for more precise estimation of effect sizes.

Characterisation of comorbid or end-organ disease in primary care EHR data sources is also limited due to missing information. For example, cirrhosis is inferred via biopsy, hepatic imaging or laboratory scores (65). However, these data are usually collected in secondary/tertiary care and are not reliably transferred into primary care EHRs. Although we identified a subset of individuals with a diagnostic code indicative of cirrhosis, cirrhotic prevalence in CHB was likely underestimated, and due to this missingness we were unable to investigate risk factors for cirrhosis as an endpoint. Similarly, the prevalence of comorbidities, including renal and cerebrovascular disease, is likely underestimated for this reason, and we are underpowered to detect associations with HCC risk. A high proportion of data on alcohol consumption is missing, and where alcohol intake is recorded, this may not be reliable. Although alcohol may be a relevant risk factor for HCC development, we are unable to report this association in our study.

## Conclusions

The CHB population in the England is ethnically diverse and socioeconomically deprived. We have identified risk factors for HCC, and validated associations observed previously in different CHB populations. Future analysis warrants validation and/or modification of existing CHB-HCC risk scores in diverse patient samples. This may require establishment of a prospective cohort study with long-term follow-up of CHB-individuals sampled from a diverse population. Missingness in various data fields an important observation: it is a limitation both for identification of individuals with CHB, and for robust description of those individuals whose diagnosis is recorded. Enhanced capture of such data is crucial to provide an evidence base for interventions, including diagnostic screening, treatment and surveillance, modification of risk factors for HCC, and monitoring progress towards elimination targets.

## Data Availability

To guarantee the confidentiality of personal and health information only the authors have had access to the data during the study in accordance with the relevant licence agreements. Access to the QResearch data is according to the information on the QResearch website (
https://www.qresearch.org/).

## ACKNOWLEDGEMENTS

We would like to thank Professor Julia Hippisley-Cox and Rebekah Burrow in the Nuffield Department of Primary Care Health Sciences at the University of Oxford for their founding/direction of and facilitation of access to the QResearch database (respectively), and for their helpful feedback on analysis. We acknowledge the contribution of EMIS practices who contribute to QResearch® and EMIS Health and the Chancellor Masters & Scholars of the University of Oxford for expertise in establishing, developing and supporting the QResearch database. QRESEARCH® is a registered trademark of Egton Medical Information Systems Limited and the University of Nottingham. This project involves data derived from patient-level information collected by the NHS, as part of the care and support of cancer patients. The data is collated, maintained and quality assured by the National Cancer Registration and Analysis Service, which is part of Public Health England (PHE). Access to the data was facilitated by the PHE Office for Data Release. This publication is based on data derived from the Intensive Care National Audit & Research Centre (ICNARC) Case Mix Programme Database. The Case Mix Programme is the national, comparative audit of patient outcomes from adult critical care coordinated by ICNARC. We thank all the staff in the critical care units participating in the Case Mix Programme. For more information on the representativeness and quality of these data, please contact ICNARC. Disclaimer: The views and opinions expressed therein are those of the authors and do not necessarily reflect those of ICNARC. The Office of National Statistics, Public Health England and NHS Digital bear no responsibility for the analysis or interpretation of the data.

## List of abbreviations

Abbreviation: Definition

95CI: 95% confidence interval

aHR: Adjusted Hazards Ration

ALT: Alanine aminotransferase

AST: Aspartate transaminase

BMI: Body mass index

CHB: Chronic hepatitis B virus infection

EHR: Electronic health record

HBsAg: Hepatitis B surface antigen

HBV: Hepatitis B virus

HCC: Hepatocellular carcinoma

HES: Hospital episode statistics

HIC: Health Informatics Collaborative

ICD: International Classification of Disease

IQR: Interquartile range

MICE: Multiple imputation by chained equations

NA: Nucleoside analogue

NCRAS: National Cancer Registration Analysis Service

NSAID: Non-steroidal anti-inflammatory drug

ONS: Office for National Statistics

Plt: Platelet

PPI: Proton pump inhibitor

SD: Standard deviation

SNOMED: Systemised Nomenclature of Medicine

T2DM: Type 2 diabetes mellitus

UK: United Kingdom

VL: Viral load

WHO: World Health Organisation

## Accessibility of protocol, raw data, and programming code

Further details regarding the study protocol are available on the study webpage (27), and a full protocol can be made available upon request to the corresponding author. Researchers interested in utilising raw QResearch data are encouraged to seek further guidance on the QResearch webpage (66). Programming code can be made available upon request to the corresponding author.

