## Supplement for "Analysis of electronic health record data of hepatitis B virus (HBV) patients in primary care: hepatocellular carcinoma (HCC) risk associated with socioeconomic deprivation and reduced by statins"

**SUPPL MATERIAL**

**Analysis of electronic health records in England of hepatitis B virus patients reveals novel risk factors for HCC including ethnicity and deprivation.**

Cori Campbell, Tingyan Wang, Iain Gillespie, Eleanor Barnes, Philippa C Matthews

**Supplementary figures and tables**

**Supplementary Figure 1**. Study flowchart depicting identification of adults from the QResearch (QR) primary care database eligible for inclusion an analysis of chronic hepatitis B (CHB) infection.

HBV DNA = hepatitis B virus DNA viral load; HBsAg = hepatitis B surface antigen; ICD = international classification of disease.

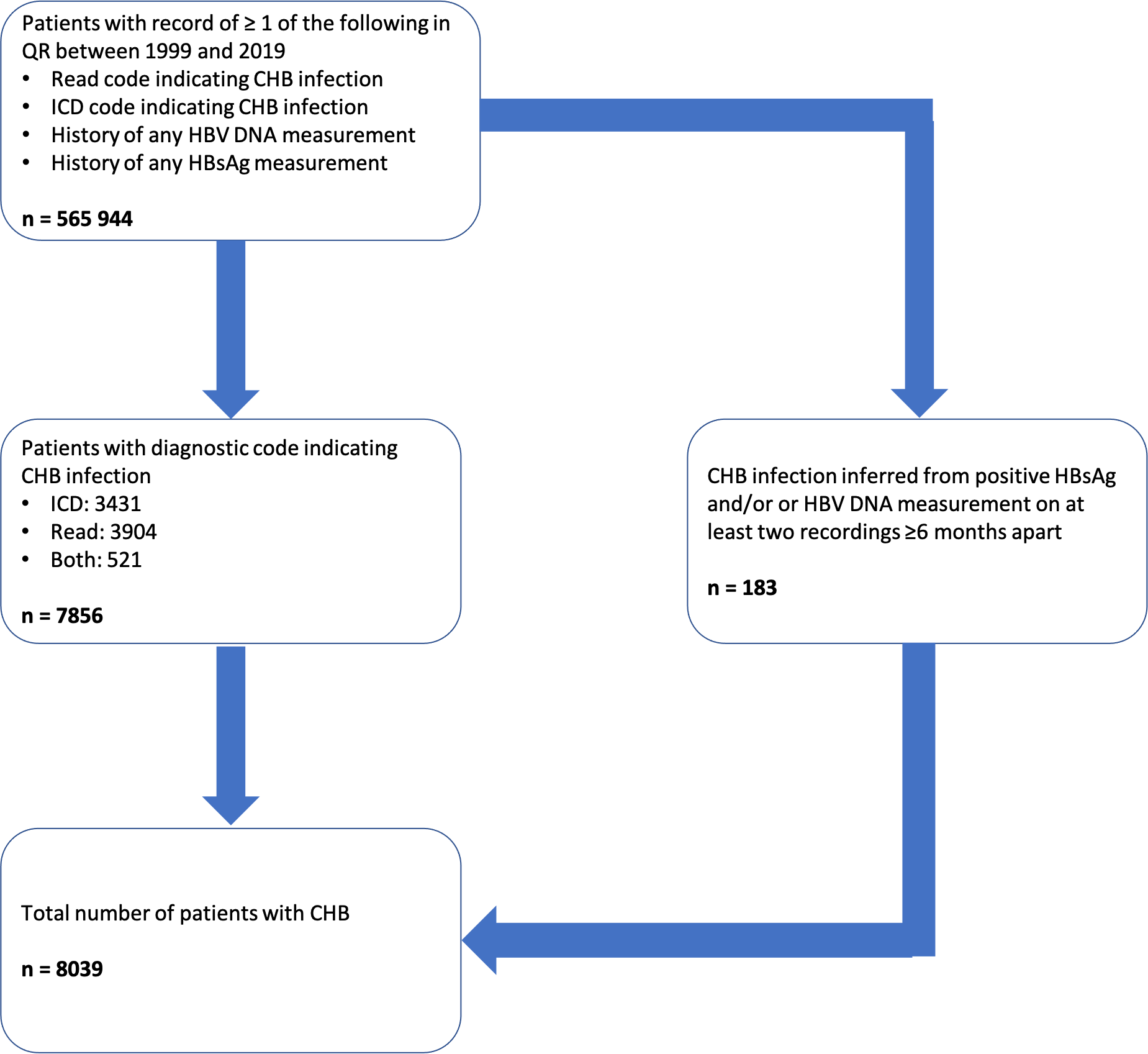

**Supplementary Table 1**. Systemised Nomenclature of Medicine primary care codes indicative of hepatocellular carcinoma used to capture cases of hepatocellular carcinoma (HCC) in adults with CHB from the QResearch database.

| **SNOMED ConceptID** | **SNOMED Fully Specified Name** |
| --- | --- |
| 92644006 | Carcinoma in situ of liver (disorder) |
| 93870000 | Malignant neoplasm of liver (disorder) |
| 95214007 | Primary malignant neoplasm of liver (disorder) |
| 109841003 | Liver cell carcinoma (disorder) |
| 187767006 | Malignant neoplasm of liver and intrahepatic bile ducts (disorder) |
| 187769009 | Primary carcinoma of liver (disorder) |
| 271525004 | Carcinoma in situ of liver and/or biliary system (disorder) |
| 395099008 | Cancer confirmed (situation) |
| 620441000000107 | Malignant neoplasm of liver and intrahepatic bile ducts NOS (disorder) |

SNOMED, Systemised Nomenclature of Medicine.

**Supplementary Table 2**. Hepatocellular carcinoma cases stratified by source of diagnosis.

| **Source of HCC diagnosis** | **Number of cases** |
| --- | --- |
| ICD | 20 |
| NCR | 22 |
| SNOMED/Read | 24 |
| ICD and NCR | 41 |
| ICD and SNOMED/Read | 15 |
| NCR and SNOMED/Read | 13 |
| ICD and NCR and SNOMED/Read | 75 |

HCC, hepatocellular carcinoma; ICD, international classification of disease; NCR, National Cancer Registry; SNOMED, Systemised Nomenclature of Medicine.

**Supplementary Table 3**. Follow-up time, stratified by source of chronic hepatitis B virus infection diagnosis.

| **Source of CHB diagnosis** | **Follow-up time (years), mean (SD) or median (IQR)*** | **CHB diagnosis date precedes cohort exit date OR HCC diagnosis date, n** | **Cohort exit date precedes CHB diagnosis date, n** | **HCC diagnosis date precedes CHB diagnosis date, n** | **Cohort exit date equates to CHB diagnosis date, n** | **HCC diagnosis date equates to CHB diagnosis date, n** |
| --- | --- | --- | --- | --- | --- | --- |
| Total | 5.12 (4.9) | 7029 | 960 | 44 | <5 | 5 |
| ICD | 7.0 (4.9) | 2430 | 958 | 29 | 0 | <5 |
| ICD and laboratory test | 7.1 (3.0)† | 12 | 0 | 0 | 0 | 0 |
| ICD and SNOMED/Read | 8.8 (7.3) | 516 | 0 | <5 | 0 | <5 |
| Laboratory test | 5.8 (2.5) | 183 | 0 | 0 | 0 | 0 |
| SNOMED/Read code | 5.0 (5.6) | 3831 | ≤5 | 11 | <5 | <5 |
| SNOMED/Read code and laboratory test | 8.7 (6.2) | 57 | 0 | 0 | 0 | 0 |

SD, standard deviation; IQR. Interquartile range; CHB, chronic hepatitis b virus; HCC, hepatocellular carcinoma; ICD, International classification of disease; SNOMED, Systemised Nomenclature of Medicine.

*Mean and SD or median and IQR calculated for patients for whom CHB diagnosis date precedes cohort exit date OR HCC diagnosis date

†Median and IQR presented

**Supplementary Table 5.** Quintiles for natural log transformed alanine transaminase (ALT), natural log transformed aspartate transaminase (AST) and platelet count (Plt). ALT and AST were natural log transformed due to their skewed distributions.

|  | **Quintile** | | | | |
| --- | --- | --- | --- | --- | --- |
| **Characteristic** | **First** | **Second** | **Third** | **Fourth** | **Fifth** |
| Log ALT |  |  |  |  |  |
| N (%) | 1429 | 1416 | 1400 | 1408 | 1376 |
| Mean log ALT (SD) | 2.69 (0.23) | 3.12 (0.09) | 3.45 (0.10) | 3.85 (0.14) | 4.79 (0.64) |
| Univariable HR (95% CI) | 0.89 (0.50 to 1.60) | 1.24 (0.73 to 2.11) | 1.00 (ref) | 1.19 (0.70 to 2.04) | 2.17 (1.35 to 3.49)** |
| Log AST |  |  |  |  |  |
| N (%) | 1378 | 1415 | 1420 | 1409 | 1407 |
| Mean log AST (SD) | 2.89 (0.14) | 3.18 (0.07) | 3.39 (0.06) | 3.67 (1.12) | 4.55 (0.69) |
| Univariable HR (95% CI) | 1.11 (0.66 to 1.88) | 0.95 (0.55 to 1.65) | 1.00 (ref) | 1.16 (0.69 to 1.96) | 1.96 (1.23 to 3.12)** |
| Plt |  |  |  |  |  |
| N (%) | 1410 | 1402 | 1393 | 1422 | 1402 |
| Mean PL (SD) | 4.80 (0.37) | 5.21 (0.05) | 5.37 (0.04) | 5.52 (0.05) | 5.76 (0.15) |
| Univariable HR (95% CI) | 4.18 (2.50 to 7.00)** | 1.18 (0.63 to 2.22) | 1.00 (ref) | 1.18 (0.63 to 2.21) | 1.38 (0.75 to 2.54) |

ALT, alanine aminotransferase; SD, standard deviation; HR, hazards ratio; CI, confidence intervals; AST, aspartate aminotransferase; Plt, platelets.

* *P* < 0.05

** *P* < 0.01

**Supplementary Table 5**. Missingness in CHB data in QResearch cohort of adults according to cohort characteristics.

| **Characteristic** | **Missingness, n (%)** |
| --- | --- |
| Age | 0 (0%) |
| Sex | 0 (0%) |
| Townsend Deprivation Quintile | 22 (0.3%) |
| Ethnicity | 1289 (16.0%) |
| Alcohol consumption | 3527 (43.9%) |
| Cigarette consumption | 2287 (28.5%) |
| BMI | 3111 (38.7%) |
| ALT | 3497 (43.5%) |
| AST | 6370 (79.2%) |
| PL | 3217 (40.0%) |
| HBsAg | 7964 (99.1%) |
| VL | 7656 (95.2%) |

BMI, body mass index; ALT, alanine transaminase; AST, aspartate transaminase; PL, platelets; HBsAg, hepatitis B virus surface antigen; VL, hepatitis B virus viral load.

**Supplementary Table 6**. Details of main and sensitivity analyses performed to investigate risk factors for hepatocellular carcinoma in QResearch cohort of adults with chronic hepatitis B virus (CHB) infection.

| Analysis | Main or sensitivity | Variables included | Corresponding table(s)/figure |
| --- | --- | --- | --- |
| Cox proportional hazards model on imputed dataset | Main | Age,  Alcohol consumption,  Antidiabetic drug use,  Antihypertensive use,  Antiviral treatment,  ARLD,  Ascites,  BMI,  Cerebrovascular disease,  CHF,  Cigarette consumption,  Cirrhosis,  CKD,  ESLD,  Ethnicity,  Hypertension,  IHD,  NAFLD,  Non-HCC neoplasm;  NSAID use,  Peptic ulcer,  Sex,  Statin use,  T2DM,  Townsend deprivation quintile, | Figure 1, Supplementary table 8 |
| Cox proportional hazards model on complete-case dataset | Sensitivity | Age,  Alcohol consumption,  Antidiabetic drug use,  Antihypertensive use,  Antiviral treatment,  ARLD,  Ascites,  BMI,  Cerebrovascular disease,  CHF,  Cigarette consumption,  Cirrhosis,  CKD,  ESLD,  Ethnicity,  Hypertension,  IHD,  NAFLD,  Non-HCC neoplasm;  NSAID use,  Peptic ulcer,  Sex,  Statin use,  T2DM,  Townsend deprivation quintile, | Supplementary table 10 |
| Cox proportional hazards model on imputed dataset whereby individuals with history of non-HCC neoplasm were excluded | Sensitivity | Age,  Alcohol consumption,  Antidiabetic drug use,  Antihypertensive use,  Antiviral treatment,  ARLD,  Ascites,  BMI,  Cerebrovascular disease,  CHF,  Cigarette consumption,  Cirrhosis,  CKD,  ESLD,  Ethnicity,  Hypertension,  IHD,  NAFLD,  NSAID use,  Peptic ulcer,  Sex,  Statin use,  T2DM,  Townsend deprivation quintile, | Supplementary table 8 |
| Cox proportional hazards model on imputed dataset with AST, ALT and Plt additionally added to model | Sensitivity | Age,  Alcohol consumption,  ALT,  Antidiabetic drug use,  Antihypertensive use,  Antiviral treatment,  ARLD,  Ascites,  AST,  BMI,  Cerebrovascular disease,  CHF,  Cigarette consumption,  Cirrhosis,  CKD,  ESLD,  Ethnicity,  Hypertension,  IHD,  NAFLD,  Non-HCC neoplasm;  NSAID use,  Peptic ulcer,  Plt,  Sex,  Statin use,  T2DM,  Townsend deprivation quintile, | Supplementary table 8 |

BMI, body mass index; T2DM, type 2 diabetes mellitus; CHF, congestive heart failure; CKD, chronic kidney disease; IHD, ischaemic heart disease; ARLD, alcohol-related liver disease; ESLD, end-stage liver disease; NAFLD, non-alcoholic fatty liver disease; NSAID, non-steroidal anti-inflammatory drug; AST, aspartate transaminase; ALT, alanine transaminase; Plt, platelets.

**Supplementary Table 7**. Tabulation of non-hepatocellular neoplasms in the cohort, overall and stratified by hepatocellular carcinoma (HCC) status.

| Type of Non-HCC neoplasm, n (%) | Overall | Non-HCC patients | HCC patients |
| --- | --- | --- | --- |
| Total | 381 (4.7 %) | 345 (4.4%) | 36 (17.1%) |
| Bone, connective tissue, skin and breast | 124 (1.5%) | 115 (1.5%) | 9 (4.3%) |
| Digestive organs and peritoneum | 44 (0.5%) | 31 (0.4%) | 13 (6.2%) |
| Eye, brain, meninges and other parts of the CNS | <5 | <5 | <5 |
| Genitourinary organs | 91 (1.1%) | 81 (1.0%) | 10 (4.8%) |
| Lip, oral cavity and pharynx | 16 (0.2%) | 16 (0.2%) | <5 |
| Lymphatic and haemopoietic tissue | 81 (1.0%) | 79 (1.0%) | <5 |
| Respiratory tract and intrathoracic organs | 13 (0.2%) | 11 (0.1%) | <5 |
| Thyroid gland and other endocrine gland structures | 8 (0.1%) | 8 (0.1%) | <5 |

HCC, hepatocellular carcinoma; CNS, central nervous system.

**Supplementary Table 8**. Cox proportional hazards model using complete imputed dataset generated by multiple imputation with chained equations (n = 7029, HCC cases = 161).

| Characteristic | Univariable HR (95% CI) | Multivariable HR (95% CI) | Multivariable HR sensitivity analysis I (95% CI) † | Multivariable HR sensitivity analysis II (95% CI) ‡ |
| --- | --- | --- | --- | --- |
| Age group, n (%) |  |  |  |  |
| 18-25 years | 0.47 (0.11 to 2.05) | 0.47 (0.11 to 2.07) | 0.45 (0.1 to 1.96) | 0.58 (0.13 to 2.7) |
| 26-35 years | 1.00 (ref) | 1.00 (ref) | 1.00 (ref) | 1.00 (ref) |
| 36-45 years | 2.49 (1.41 to 4.38)** | 1.96 (1.1 to 3.48)* | 1.82 (1.01 to 3.27)* | 1.83 (1.01 to 3.3)* |
| 46-55 years | 5.26 (3.02 to 9.15)** | 2.73 (1.51 to 4.95)** | 2.55 (1.38 to 4.72)** | 2.2 (1.21 to 4.01)* |
| 56-65 years | 7.52 (4.14 to 13.67)** | 3.17 (1.63 to 6.14)** | 3.49 (1.77 to 6.89)** | 2.26 (1.13 to 4.54)* |
| ≥66 years | 11.89 (6.26 to 22.60)** | 4.06 (1.92 to 8.58)** | 3.3 (1.42 to 7.72)** | 2.41 (1.12 to 5.19)* |
| Sex, n (%) |  |  |  |  |
| Female | 1.00 (ref) | 1.00 (ref) | 1.00 (ref) | 1.00 (ref) |
| Male | 4.92 (3.05 to 7.95)** | 3.44 (2.07 to 5.73)** | 3.61 (2.05 to 6.34)** | 3.79 (2.25 to 6.36)** |
| Townsend Deprivation Quintile, n (%) |  |  |  |  |
| First (least deprived) | 2.06 (1.09 to 3.9)* | 1.87 (0.96 to 3.65) | 1.85 (0.89 to 3.82) | 1.84 (0.92 to 3.67) |
| Second | 1.15 (0.59 to 2.24) | 1.1 (0.54 to 2.25) | 1.07 (0.49 to 2.32) | 1.16 (0.55 to 2.43) |
| Third | 1.00 (ref) | 1.00 (ref) | 1.00 (ref) | 1.00 (ref) |
| Fourth | 1.11 (0.64 to 1.93) | 1.27 (0.72 to 2.24) | 1.11 (0.59 to 2.07) | 1.23 (0.68 to 2.24) |
| Fifth (most deprived) | 1.34 (0.82 to 2.19) | 1.69 (1.01 to 2.84)* | 1.66 (0.96 to 2.88) | 1.63 (0.95 to 2.8) |
| Ethnicity, n (%) |  |  |  |  |
| White | 1.00 (ref) | 1.00 (ref) | 1.00 (ref) |  |
| Black African | 0.74 (0.48 to 1.14) | 0.99 (0.61 to 1.61) | 1.07 (0.64 to 1.8) | 1.00 (ref) |
| Bangladeshi | 1.29 (0.67 to 2.47) | 1.05 (0.52 to 2.14) | 1.14 (0.55 to 2.37) | 1.09 (0.53 to 2.22) |
| Caribbean | 2.23 (1.01 to 4.89)* | 3.32 (1.43 to 7.71)** | 4.26 (1.73 to 10.52)** | 1.2 (0.43 to 3.34) |
| Chinese | 0.75 (0.44 to 1.28) | 0.85 (0.47 to 1.52) | 0.97 (0.53 to 1.77) | 0.59 (0.32 to 1.09) |
| Indian | 1.67 (0.82 to 3.38) | 1.83 (0.84 to 3.98) | 1.91 (0.83 to 4.41) | 1.29 (0.53 to 3.12) |
| Other | 0.73 (0.42 to 1.28) | 1.06 (0.59 to 1.89) | 0.75 (0.37 to 1.52) | 0.76 (0.41 to 1.41) |
| Other Asian | 0.59 (0.25 to 1.37) | 0.87 (0.36 to 2.08) | 1.03 (0.42 to 2.52) | 0.61 (0.27 to 1.39) |
| Pakistani | 0.47 (0.2 to 1.1) | 0.67 (0.27 to 1.66) | 0.76 (0.29 to 1.98) | 0.68 (0.3 to 1.51) |
| Cigarette consumption, n (%) |  |  |  |  |
| Non-smoker | 1.00 (ref) | 1.00 (ref) | 1.00 (ref) | 1.00 (ref) |
| Ex-smoker | 1.1 (0.68 to 1.78) | 0.77 (0.46 to 1.27) | 0.89 (0.52 to 1.52) | 0.63 (0.37 to 1.07) |
| Light smoker (1-9 cigarettes/day) | 1.73 (1.19 to 2.53)** | 1.41 (0.94 to 2.12) | 1.47 (0.94 to 2.28) | 1.18 (0.76 to 1.82) |
| Moderate to heavy smoker (≥10 cigarettes per day) | 1.34 (0.67 to 2.65) | 1.05 (0.51 to 2.18) | 1.3 (0.61 to 2.78) | 1.82 (0.95 to 3.49) |
| Alcohol consumption, n (%) |  |  |  |  |
| Non-drinker | 1.00 (ref) | 1.00 (ref) | 1.00 (ref) | 1.00 (ref) |
| Trivial drinker (<1 unit per day) | 1.24 (0.78 to 1.98) | 1.4 (0.86 to 2.28) | 1.23 (0.71 to 2.14) | 1.02 (0.57 to 1.82) |
| Light drinker (1-2 units per day) | 0.81 (0.36 to 1.84) | 0.73 (0.32 to 1.69) | 0.81 (0.35 to 1.9) | 0.54 (0.2 to 1.46) |
| Moderate to heavy drinker (≥3 units per day) | 1.26 (0.52 to 3.08) | 0.87 (0.34 to 2.23) | 1.22 (0.48 to 3.09) | 0.63 (0.23 to 1.69) |
| BMI, n (%) |  |  |  |  |
| Underweight (<18.5 kg/m^2^) | 1.3 (0.56 to 3) | 1.31 (0.55 to 3.15) | 1.63 (0.64 to 4.14) | 0.65 (0.19 to 2.21) |
| Normal weight (18.5-24.9 kg/m^2^) | 1.00 (ref) | 1.00 (ref) | 1.00 (ref) | 1.00 (ref) |
| Overweight (25.0-29.9 kg/m^2^) | 1.07 (0.75 to 1.52) | 1.06 (0.74 to 1.52) | 1 (0.68 to 1.48) | 0.86 (0.59 to 1.25) |
| Obese (≥30 kg/m^2^) | 1.06 (0.68 to 1.63) | 1.02 (0.64 to 1.62) | 1 (0.6 to 1.64) | 0.79 (0.49 to 1.28) |
| Antiviral initiation, n (%) |  |  |  |  |
| No initiation | 1.00 (ref) | 1.00 (ref) | 1.00 (ref) | 1.00 (ref) |
| Before CHB diagnosis | 3.67 (1.71 to 7.9)** | 1.85 (0.79 to 4.32) | 2.15 (0.88 to 5.24) | 1.39 (0.59 to 3.3) |
| Within 1 year of CHB diagnosis | 4.93 (2.95 to 8.24)** | 1.63 (0.92 to 2.9) | 1.48 (0.8 to 2.76) | 1.66 (0.91 to 3.03) |
| Within 2 years of CHB diagnosis | 3.42 (1.59 to 7.35)** | 1.24 (0.55 to 2.81) | 1.64 (0.73 to 3.72) | 1.41 (0.61 to 3.23) |
| Within 3 years of CHB diagnosis | 4.32 (2.01 to 9.29)** | 1.89 (0.83 to 4.28) | 1.91 (0.78 to 4.7) | 2.06 (0.88 to 4.87) |
| ≥4 years after CHB diagnosis | 1.55 (0.95 to 2.55) | 0.76 (0.45 to 1.27) | 0.62 (0.34 to 1.13) | 0.49 (0.28 to 0.84) |
| Type 2 DM, n (%) | 2.57 (1.79 to 3.7)** | 1.57 (0.72 to 3.41) | 1.07 (0.43 to 2.68) | 1.46 (0.63 to 3.37) |
| Congestive heart failure, n (%) | 2.1 (0.78 to 5.68) | 0.77 (0.27 to 2.15) | 1.03 (0.36 to 2.96) | 0.7 (0.24 to 2.07) |
| Hypertension, n (%) | 2.32 (1.68 to 3.22)** | 1.19 (0.81 to 1.74) | 1.15 (0.76 to 1.76) | 0.94 (0.63 to 1.4) |
| Chronic kidney disease, n (%) | 3.83 (2.5 to 5.88)** | 1.26 (0.76 to 2.08) | 1.31 (0.74 to 2.32) | 1.01 (0.61 to 1.68) |
| Alcohol-related liver disease, n (%) | 5.82 (3.06 to 11.05)** | 0.95 (0.45 to 2.01) | 1.17 (0.54 to 2.49) | 1 (0.45 to 2.22) |
| Ascites, n (%) | 13.61 (8.42 to 21.99)** | 1.85 (1.02 to 3.36)* | 2.08 (1.07 to 4.05)* | 1.68 (0.93 to 3.07) |
| Cerebrovascular disease, n (%) | 2.31 (1.4 to 3.83)** | 0.64 (0.3 to 1.36) | 0.72 (0.32 to 1.64) | 0.65 (0.3 to 1.41) |
| Cirrhosis, n (%) | 12.65 (9.22 to 17.36)** | 6.52 (4.54 to 9.37)** | 7.52 (5.07 to 11.14)** | 5.11 (3.5 to 7.47)** |
| End-stage liver disease, n (%) | 7.66 (4.34 to 13.52)** | 1.24 (0.62 to 2.47) | 0.92 (0.41 to 2.06) | 1.75 (0.88 to 3.45) |
| Ischaemic heart disease, n (%) | 3.01 (1.82 to 4.99)** | 1.9 (0.88 to 4.11) | 1.69 (0.7 to 4.08) | 1.67 (0.75 to 3.73) |
| Non-alcoholic fatty liver disease, n (%) | 0.69 (0.32 to 1.48) | 0.66 (0.3 to 1.46) | 0.52 (0.21 to 1.31) | 0.75 (0.33 to 1.68) |
| Non-HCC neoplasm, n (%) | 2.3 (1.45 to 3.67)** | 0.95 (0.57 to 1.58) | -- |  |
| Peptic ulcer, n (%) | 5.54 (3.68 to 8.34)** | 2.20 (1.39 to 3.49)** | 2.33 (1.42 to 3.82)** | 1.96 (1.22 to 3.15) |
| Antidiabetic drug use, n (%) | 2.53 (1.75 to 3.65)** | 1.83 (0.74 to 4.56) | 1.98 (0.73 to 5.37) | 1.09 (0.4 to 2.94) |
| Antihypertensive use, n (%) | 2.2 (1.39 to 3.48)** | 0.48 (0.21 to 1.07) | 0.48 (0.19 to 1.23) | 0.92 (0.4 to 2.15) |
| NSAID use, n (%) | 1.96 (1.24 to 3.1)** | 0.96 (0.48 to 1.96) | 0.96 (0.44 to 2.09) | 1.16 (0.54 to 2.49) |
| Statin use, n (%) | 1.72 (1.06 to 2.78)* | 0.47 (0.22 to 0.99)* | 0.64 (0.26 to 1.56) | 0.38 (0.17 to 0.84) |
| ALT quintile, n (%) |  |  |  |  |
| First quintile (mean log ALT=, SD=) | -- | -- | -- | 0.95 (0.46 to 1.99) |
| Second quintile (mean log ALT=, SD=) | -- | -- | -- | 1.91 (1.03 to 3.56)* |
| Third quintile (mean log ALT=, SD=) | -- | -- | -- | 1.00 (ref) |
| Fourth quintile (mean log ALT=, SD=) | -- | -- | -- | 1.93 (1.08 to 3.46)* |
| Fifth quintile (mean log ALT=, SD=) | -- | -- | -- | 2.34 (1.35 to 4.06)** |
| AST quintile, n (%) |  |  |  |  |
| First quintile (mean log AST=, SD=) | -- | -- | -- | 0.6 (0.33 to 1.07) |
| Second quintile (mean log AST=, SD=) | -- | -- | -- | 0.99 (0.56 to 1.73) |
| Third quintile (mean log AST=, SD=) | -- | -- | -- | 1.00 (ref) |
| Fourth quintile (mean log AST=, SD=) | -- | -- | -- | 0.84 (0.5 to 1.4) |
| Fifth quintile (mean log AST=, SD=) | -- | -- | -- | 0.97 (0.6 to 1.57) |
| PL quintile, m (%) |  |  |  |  |
| First quintile (mean log PL=, SD=) | -- | -- | -- | 1.25 (0.75 to 2.08) |
| Second quintile (mean log PL=, SD=) | -- | -- | -- | 0.67 (0.36 to 1.23) |
| Third quintile (mean log PL=, SD=) | -- | -- | -- | 1.00 (ref) |
| Fourth quintile (mean log PL=, SD=) | -- | -- | -- | 0.79 (0.41 to 1.52) |
| Fifth quintile (mean log PL=, SD=) | -- | -- | -- | 1.01 (0.56 to 1.84) |

HR, hazards ratio; CHB, chronic hepatitis B virus infection; ICD, International classification of disease; SNOMED, Systemised Nomenclature of Medicine; SD, standard deviation; BMI, body mass index; DM, diabetes mellitus; HCC, hepatocellular carcinoma; NSAID, non-steroidal anti-inflammatory drug.

* *P* < 0.05

** *P* < 0.01

† Patients with history of any non-hepatocellular carcinoma neoplasm excluded.

‡ AST, ALT and Plt added to model.

**Supplementary Table 9**. Cox proportional hazards complete-case model conducted on adults with hepatitis B virus infection for whom data was complete in the original QResearch dataset (n = 3648, HCC cases = 68).

| Characteristic | Univariable HR (95% CI) | Multivariable HR (95% CI) | Multivariable HR (95% CI) † |
| --- | --- | --- | --- |
| Age group, n (%) |  |  |  |
| 18-25 years | 0.85 (0.19 to 3.88) | 1.7 (0.35 to 8.26) | 1.98 (0.4 to 9.91) |
| 26-35 years | 1.00 (ref) | 1.00 (ref) | 1.00 (ref) |
| 36-45 years | 1.59 (0.72 to 3.51) | 1.44 (0.61 to 3.37) | 1.46 (0.61 to 3.48) |
| 46-55 years | 3.14 (1.43 to 6.85)** | 2.4 (0.99 to 5.82) | 1.91 (0.75 to 4.85) |
| 56-65 years | 6.84 (3.13 to 14.97)** | 3.24 (1.23 to 8.53)** | 3.89 (1.43 to 10.61)** |
| ≥66 years | 5.47 (1.98 to 15.14)** | 2.41 (0.7 to 8.37) | 1.7 (0.37 to 7.73) |
| Sex, n (%) |  |  |  |
| Female | 1.00 (ref) | 1.00 (ref) | 1.00 (ref) |
| Male | 5.50 (2.63 to 11.50)** | 3.35 (1.42 to 7.88)** | 3.49 (1.35 to 8.99)** |
| Townsend Deprivation Quintile, n (%) |  |  |  |
| First (least deprived) | 3.12 (0.95 to 10.24) | 3.6 (1 to 12.99) | 3.95 (0.9 to 17.35) |
| Second | 1.91 (0.55 to 6.61) | 2.47 (0.63 to 9.74) | 1.32 (0.27 to 6.54) |
| Third | 1.00 (ref) | 1.00 (ref) | 1.00 (ref) |
| Fourth | 1.74 (0.63 to 4.79) | 1.69 (0.57 to 5.02) | 1.78 (0.55 to 5.74) |
| Fifth (most deprived) | 2.18 (0.86 to 5.56) | 2.34 (0.84 to 6.53) | 2.75 (0.92 to 8.16) |
| Ethnicity, n (%) |  |  |  |
| White | 1.00 (ref) | 1.00 (ref) | 1.00 (ref) |
| Black African | 1.14 (0.58 to 2.24) | 2.37 (0.96 to 5.81) | 2.95 (1.13 to 7.73)* |
| Bangladeshi | 2.16 (0.92 to 5.05) | 1.77 (0.63 to 4.96) | 1.88 (0.64 to 5.55) |
| Caribbean | 1.91 (0.44 to 8.32) | 3.32 (0.59 to 18.6) | 8.31 (1.59 to 43.59)* |
| Chinese | 0.68 (0.25 to 1.85) | 1.3 (0.42 to 4.02) | 2 (0.6 to 6.6) |
| Indian | 2.52 (0.92 to 6.88) | 5.19 (1.57 to 17.09)** | 7.23 (1.98 to 26.49)* |
| Other | 1.14 (0.49 to 2.66) | 2.18 (0.8 to 5.92) | 1.41 (0.43 to 4.67) |
| Other Asian | 0.6 (0.14 to 2.63) | 1.41 (0.28 to 7.03) | 1.49 (0.28 to 8.1) |
| Pakistani | 0.86 (0.29 to 2.57) | 1.87 (0.53 to 6.61) | 1.82 (0.48 to 6.84) |
| Cigarette consumption, n (%) |  |  |  |
| Non-smoker | 1.00 (ref) | 1.00 (ref) | 1.00 (ref) |
| Ex-smoker | 1.57 (0.77 to 3.17) | 1.1 (0.51 to 2.39) | 1.56 (0.68 to 3.56) |
| Light smoker (1-9 cigarettes/day) | 2.91 (1.68 to 5.06)** | 2.29 (1.15 to 4.57)* | 3.28 (1.54 to 6.97)** |
| Moderate to heavy smoker (≥10 cigarettes per day) | 1.96 (0.70 to 5.53) | 2.92 (0.92 to 9.29) | 3.52 (1.06 to 11.67)* |
| Alcohol consumption, n (%) |  |  |  |
| Non-drinker | 1.00 (ref) | 1.00 (ref) | 1.00 (ref) |
| Trivial drinker (<1 unit per day) | 1.17 (0.56 to 2.45) | 1.79 (0.79 to 4.02) | 1.48 (0.59 to 3.71) |
| Light drinker (1-2 units per day) | 0.66 (0.16 to 2.71) | 0.45 (0.1 to 2.13) | 0.6 (0.13 to 2.78) |
| Moderate to heavy drinker (≥3 units per day) | 1.25 (0.31 to 5.14) | 0.46 (0.09 to 2.3) | 1.15 (0.22 to 5.93) |
| BMI, n (%) |  |  |  |
| Underweight (<18.5 kg/m^2^) | 0.42 (0.06 to 3.11) | 0.95 (0.12 to 7.78) | 1.74 (0.21 to 14.22) |
| Normal weight (18.5-24.9 kg/m^2^) | 1.00 (ref) | 1.00 (ref) | 1.00 (ref) |
| Overweight (25.0-29.9 kg/m^2^) | 0.92 (0.55 to 1.55) | 1 (0.57 to 1.77) | 1.17 (0.62 to 2.18) |
| Obese (≥30 kg/m^2^) | 0.54 (0.26 to 1.15) | 0.68 (0.29 to 1.63) | 0.9 (0.36 to 2.25) |
| Antiviral initiation, n (%) |  |  |  |
| No initiation | 1.00 (ref) | 1.00 (ref) | 1.00 (ref) |
| Before CHB diagnosis | 3.29 (1.01 to 10.68)* | 1.78 (0.45 to 6.97) | 2.87 (0.71 to 11.65) |
| Within 1 year of CHB diagnosis | 5.11 (2.45 to 10.66)** | 2.32 (1 to 5.38) | 1.84 (0.74 to 4.59) |
| Within 2 years of CHB diagnosis | 4.04 (1.57 to 10.39)** | 1.46 (0.5 to 4.26) | 1.51 (0.51 to 4.44) |
| Within 3 years of CHB diagnosis | 6.03 (2.36 to 15.37)** | 2.36 (0.82 to 6.81) | 2.26 (0.7 to 7.35) |
| ≥4 years after CHB diagnosis | 1.95 (0.90 to 4.23) | 0.92 (0.4 to 2.1) | 0.67 (0.24 to 1.87) |
| Type 2 DM, n (%) | 3.26 (1.96 to 5.42)** | 2.5 (0.75 to 8.34) | 1.74 (0.42 to 7.32) |
| Congestive heart failure, n (%) | 1.06 (0.15 to 7.70) | 0.22 (0.03 to 1.92) | 0.19 (0.02 to 1.82) |
| Hypertension, n (%) | 2.29 (1.41 to 3.73) | 1.05 (0.56 to 1.96) | 1.11 (0.56 to 2.21) |
| Chronic kidney disease, n (%) | 5.48 (3.14 to 9.53)** | 3.32 (1.65 to 6.70)** | 3.11 (1.43 to 6.79)** |
| Alcohol-related liver disease, n (%) | 5.79 (2.50 to 13.42)** | 1.51 (0.49 to 4.64) | 1.18 (0.37 to 3.72) |
| Ascites, n (%) | 18.11 (8.63 to 37.99)** | 1.81 (0.69 to 4.74) | 1.51 (0.55 to 4.17) |
| Cerebrovascular disease, n (%) | 2.35 (1.16 to 4.76)* | 1.08 (0.37 to 3.20)** | 1.48 (0.46 to 4.72) |
| Cirrhosis, n (%) | 17.12 (10.37 to 28.25)** | 8.79 (4.85 to 15.93)** | 11.54 (5.95 to 22.39)** |
| End-stage liver disease, n (%) | 9.28 (3.71 to 23.20)** | 1.52 (0.46 to 5.09) | 1.73 (0.46 to 6.51) |
| Ischaemic heart disease, n (%) | 1.76 (0.75 to 4.10) | 0.37 (0.1 to 1.4) | 0.31 (0.07 to 1.33) |
| Non-alcoholic fatty liver disease, n (%) | 0.60 (0.19 to 1.91) | 0.56 (0.16 to 1.93) | 0.63 (0.18 to 2.2) |
| Non-HCC neoplasm, n (%) | 2.49 (1.19 to 5.22)* | 1.23 (0.5 to 3.03) | -- |
| Peptic ulcer, n (%) | 4.21 (2.18 to 8.13)** | 2.20 (1.03 to 4.70)* | 3.07 (1.35 to 7)** |
| Antidiabetic drug use, n (%) | 3.02 (1.82 to 4.99)** | 1.43 (0.37 to 5.56) | 1.47 (0.31 to 6.95) |
| Antihypertensive use, n (%) | 2.56 (1.42 to 4.62)** | 0.6 (0.2 to 1.78) | 0.62 (0.18 to 2.17) |
| NSAID use, n (%) | 1.98 (1.08 to 3.64)* | 0.81 (0.32 to 2.09) | 0.88 (0.31 to 2.53) |
| Statin use, n (%) | 1.81 (0.97 to 3.38) | 0.67 (0.25 to 1.81) | 0.93 (0.29 to 2.98) |

HR, hazard ratio; CHB, chronic hepatitis B virus infection; ICD, International classification of disease; SNOMED, Systemised Nomenclature of Medicine; SD, standard deviation; BMI, body mass index; DM, diabetes mellitus; HCC, hepatocellular carcinoma; NSAID, non-steroidal anti-inflammatory drug.

* *P* < 0.05

** *P* < 0.01

† Patients with history of any non-hepatocellular carcinoma neoplasm excluded
